# Paired DNA and RNA sequencing uncovers common and rare genetic variants regulating gene expression in the human retina

**DOI:** 10.1101/2025.04.25.25326445

**Authors:** Jacob Sampson, Ayellet V Segrè, Kinga M Bujakowska, Steve Haynes, Diana Baralle, Siddharth Banka, Graeme C Black, Panagiotis I Sergouniotis, Jamie M Ellingford

## Abstract

Genetic disorders impacting vision affect millions of individuals worldwide, including age-related macular degeneration (common) and inherited retinal disorders (rare). There is incomplete understanding of the impact of genetic variation on gene expression in the human retina, and its role in genetic disorders. Through the generation of whole genome sequencing and bulk RNA-sequencing of neurosensory retina (NSR) and retinal pigment epithelium (RPE) from 201 post-mortem eyes, we uncovered common and rare genetic variants shaping retinal expression profiles. This includes 1,483,595 significant cis-expression quantitative trait loci (eQTLs) impacting 9,959 and 3,699 genes in NSR and RPE, respectively, with associated genetic variants enriched to cis-candidate regulatory elements and notable shared eGenes between NSR and RPE. We also detected 1051 expression outliers and prioritised 299 rare non-coding single-nucleotide, structural variants or copy number variants as plausible drivers for 28% of outlier events. This study increases understanding of gene expression regulation in the human retina.

**Graphical Abstract:** 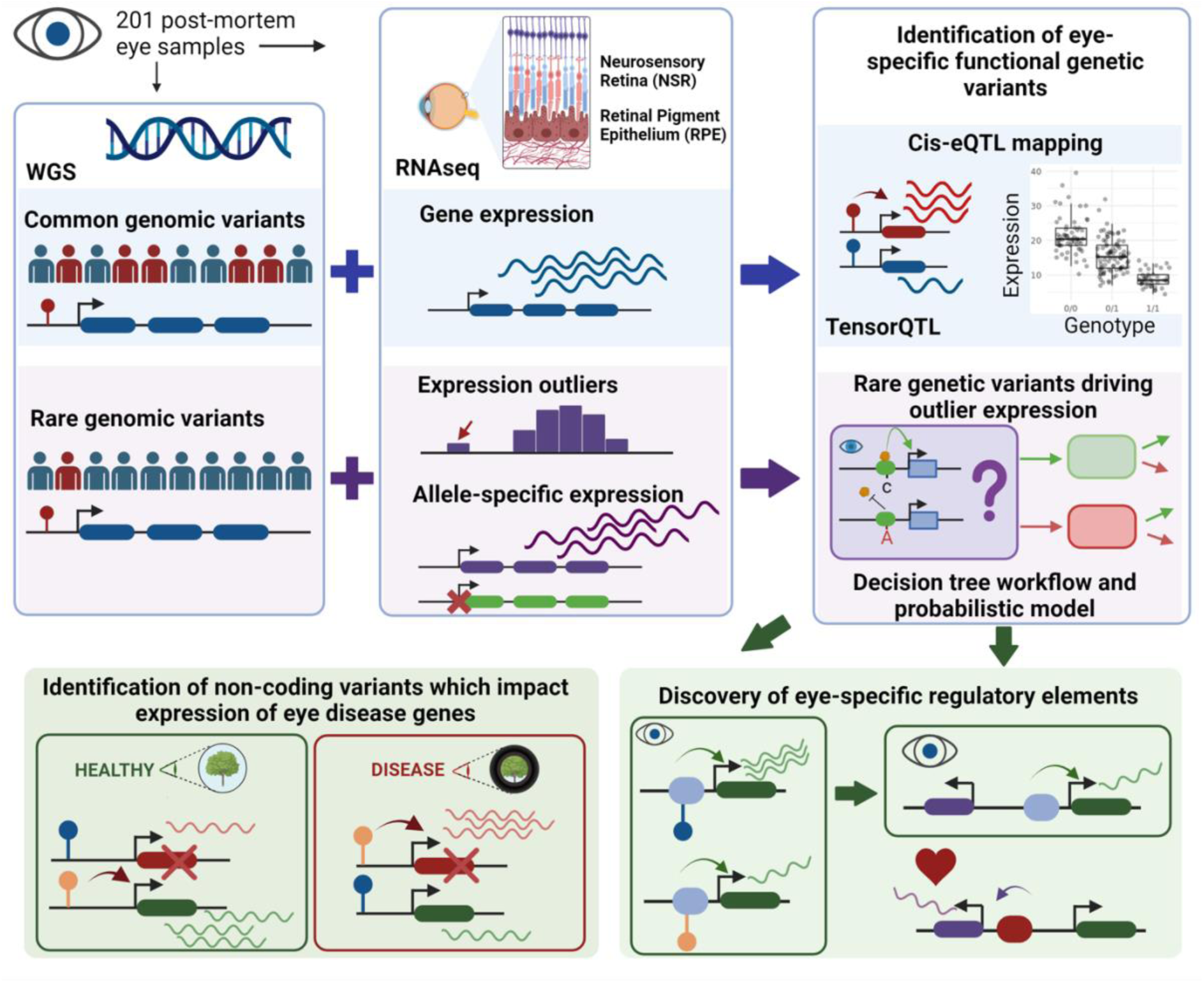

## Introduction

Genetic variation has been well established to play a role in the onset and susceptibility of visual impairment by disrupting normal functioning of the retina, a highly specialised light-sensitive tissue at the back of eye (Wright *et al*., 2010). The retina depends on the interaction between neuronal and non-neuronal cell types, including those in the neurosensory retina (NSR), e.g. photoreceptors and ganglion cells (Hoon *et al*., 2014), and the retinal pigment epithelium (RPE), a monolayer which lines the photoreceptor outer segments (Strauss, 2005). Inherited retinal disorders (IRDs) are a diverse set of largely monogenic conditions driven primarily by highly impactful genetic variants that are rare in the population and disrupt function of the NSR and/or RPE (Hanany, Rivolta and Sharon, 2020). Monogenic IRDs may present in isolation, for example, Stargardt disease, retinitis pigmentosa and cone-rod dystrophy, or as part of a multi-system disorder, for example Usher syndrome, Joubert syndrome and Senior-Loken syndrome. Age-related macular degeneration (AMD) is a common disorder that impacts the retina and is a leading cause of visual impairment in adults (Fleckenstein *et al*., 2021), predicted to impact 288 million individuals by 2040 (Wong *et al*., 2014). Whilst non-genetic risk factors exist for AMD, including age, diet and lifestyle, its heritability is estimated to be as high as 71% (Seddon *et al*., 2005). Genome wide association studies (GWAS) initially identified more than 50 genomic loci impacting 34 genes that convey high risk to AMD in a European ancestry cohort (Fritsche *et al*., 2016). Recent expansion of AMD GWAS to Hispanic and African ancestries has uncovered 30 additional genomic loci and distinct AMD genomic architecture in these populations (Gorman *et al*., 2024).

The Genotype-Tissue Expression (GTEx) project has transformed our ability to pinpoint genetic variants that impact gene expression (THE GTEX CONSORTIUM, 2020), including tissue-specific and tissue-shared expression quantitative loci (eQTLs) and rare genetic variants associated with expression outliers (eOutliers). Findings from these investigations, along with other studies, have been leveraged across various biological and medical fields to gain a deeper understanding of disease mechanisms (Barbeira *et al*., 2021; Hamel *et al*., 2024), to provide more informed diagnosis and prognosis (Michaud *et al*., 2022), and to pursue pathways for novel treatments (Davenport *et al*., 2018).

Notably, ophthalmic tissue was not included in the GTEx resource, but recently, the EyeGEx study identified over 2 million eQTLs in the retina, from a cohort comprised of healthy eyes and eyes displaying signs of AMD (Ratnapriya *et al*., 2019). This resource enables the investigation of the role of common single-nucleotide variants (SNVs) influencing retinal gene expression. However, to our knowledge, there are no suitable datasets to also interrogate the impact of rare variants, structural variants (SVs) and copy number variants (CNVs) on retinal gene expression. Expanding our understanding of gene expression regulation in the retina will provide insights into the molecular mechanisms underlying both common and rare eye diseases, and help identify new potential strategies for treatment and prevention.

Here we describe the creation of a unique resource of paired genomes and transcriptomes for the human retina from 201 donors, and develop new understanding of both common and rare variants which drive expression in this highly specialised tissue.

## Results

### The METR genome-transcriptome resource integrates genomic and retinal transcriptomic data from 201 post-mortem eye samples

The Manchester Eye Tissue Repository (METR) genome-transcriptome cohort is comprised of 201 unrelated individuals that donated eye tissue post-mortem. The median age of the cohort was 71 years (IQR 64-77) at time of post-mortem, with a slight male predominance (63.7%). The median ischemic time was 40 hours *(IQR 32-44).* While 47 individuals (23% of the cohort) were found to carry genetic variants that confer high-risk for age-related macular degeneration (AMD), none of the 201 individuals included in the cohort had phenotypic presentation, assessed post-mortem, consistent with late-stage AMD or monogenic ophthalmic disorders.

Short-read whole genome sequencing was performed on an Illumina NovaSeq6000, with alignment and variant detection performed using DRAGEN software (v4.0.3). The median genome-wide average coverage per sample was 35.9x (IQR 30.3 – 40.5), with an average of 88.0% and 92.8% of the genome covered by at least 15 or at least 10 sequencing reads, respectively. Joint SNV calling with DRAGEN PopGen 4.2.4 obtained aggregate calls at 15,617,784 high-confidence variant sites after quality control (***Supplementary table 1***). On average, 173 CNVs and 8,814 SVs were identified per sample (***Supplementary table 2***).

Transcriptomic data was generated by short read bulk**-**RNA sequencing of polyadenylated enriched RNA, using an Illumina NovaSeq6000. The median number of uniquely mapped reads for NSR samples (n = 183) was 139 million and for RPE samples (n = 176) was 62.3 million (***Supplementary figure 1***). Some level of expression (mean TPM > 0.1) was indicated for 28,512 genes across both tissues, with 18,891 and 13,214 genes expressed at moderate (TPM >1) and high (TPM > 5) levels, respectively (**Figure 1A**). Among the expressed genes, 16,765 (59%) were protein coding, representing 84% of all GENCODE protein-coding genes. Significantly higher expression variability across samples was observed in the RPE compared to the NSR for genes expressed at low, moderate and high levels (p-value<2.2×10^-16^; **Figure 1B**).

**Figure 1.**
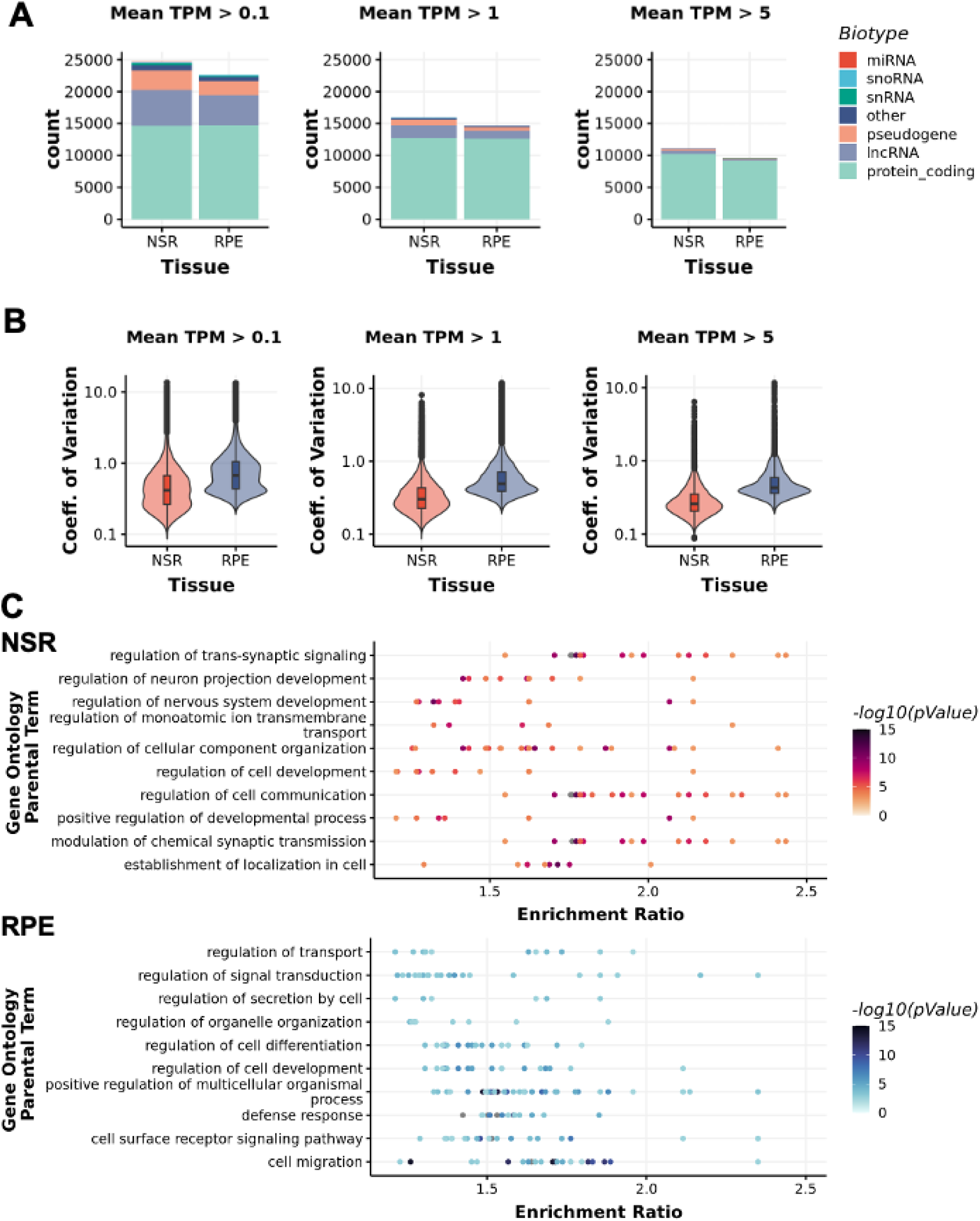
Tissue-specific transcriptomic data generated for the neurosensory retina (NSR) and the retinal pigment epithelium (RPE) A) Number of genes expressed in the NSR and the RPE al different expression thresholds, classified into different biotype’s. B) The coefficient of variation (mean/SD) for all genes expressed at different thresholds in both tissues indicates higher expression variability across samples in the RPE compared to the NSR. C) Top 10 Gene Ontology Terms enriched in differentially expressed genes (adj pvalue < 0.01) in the NSR and RPE. respectively. Gene Ontology terms (FDR < 0.01) were sorted by enrichment ratio and redundant terms were removed.

Gene expression profiles were enriched for gene ontology (GO) terms indicative of the tissues of origin (**Figure 1C**). Overall, 21,157 differentially expressed genes (adj. p-value < 0.01) were identified between the RPE and the NSR. Unsurprisingly, cell type deconvolution analyses demonstrated a significantly higher representation of RPE cells in data generated from RPE samples compared to data generated from NSR samples (**Supplementary figure 2A**). Moreover, genes with increased expression in the RPE (n = 9,606) were enriched for 969 GO terms, including epithelial cell proliferation (GO:0050678) and innate and adaptive immune responses (for example, GO:0045087; GO:0002250) (***Supplementary table 3***). Deconvolution of the NSR datasets supported the presence of at least 7 neuronal cell types at high levels (estimated proportion > 1%), with an average relative composition, per sample, of 29% rod photoreceptors (95%CI:26.4-30.7%), 28% retinal astrocytes (95%CI=26.1-29.6%), 16% amacrine cells (95%CI=15.7-17.1%), 10% horizontal cells (95%CI=9.5-10.2%), 7% retinal ganglion cells (95%CI=5.8-9%), 4% bipolar cells (95%CI=3.8-4.4%), 2% Müller glia (95%CI=1.7-3.1%) and ∼4% other cell types (**Supplementary figure 2C**). Genes with increased expression in the NSR, compared to the RPE (n = 11,551), were enriched for 237 GO terms including nervous system development (GO: 0007399) neuronal signal transduction (GO:0023041), synapse assembly (GO:0007416) and synaptic signalling (GO:0099536) (***Supplementary table 4***).

### METR eQTLs provide novel insights into non-coding variants that impact known eye disease-related genes

We performed cis-eQTL mapping to identify common genetic variants that are associated with gene expression in the NSR and the RPE. We found 1,424,946 significant (FDR < 0.05) cis-eQTL associations between 806,789 variants (eVariants) and 9,959 genes (eGenes) in NSR. Additionally, 465,045 eQTLs were identified between 303,773 eVariants and 3,699 eGenes in the RPE (**Supplementary figure 3**). 406,396 eQTLs were common to both the retina and the RPE, while 1,018,550 associations were NSR-specific and 58,649 were RPE-specific (**Figure 2A**). Henceforth, we will refer to eQTLs identified in the NSR and/or the RPE as METR-eQTLs (n=1,483,595), which included 10,471 unique eGenes (6,772 NSR-specific, 512 RPE-specific and 3,187 eGenes in both NSR and RPE).

**Figure 2.**
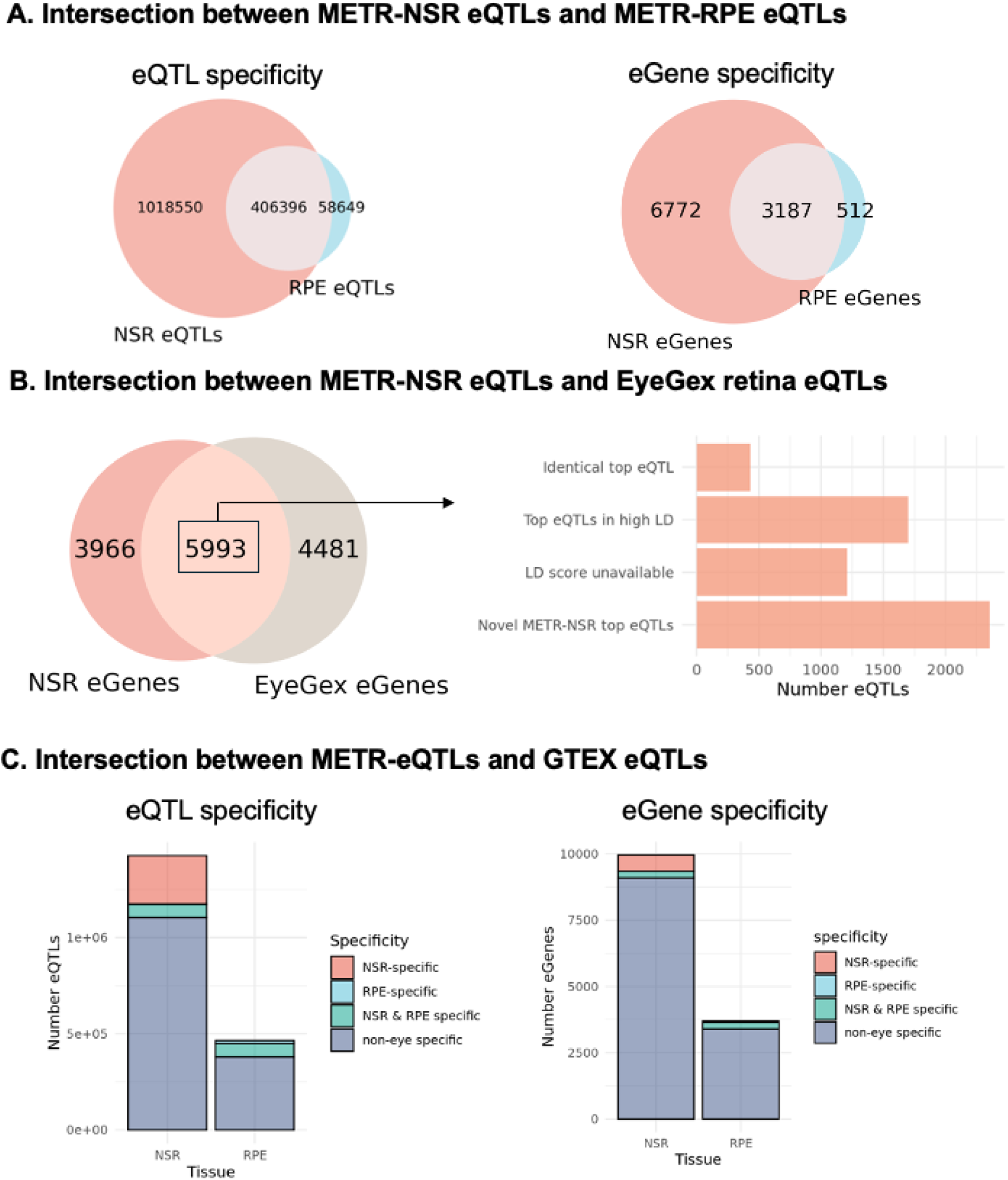
Intersection between METR-eQTls and eQTL.s from different studies. A) Overlap between METR-eQTLs (Jeff.) and METR-eGenes (right) identified in the neurosensory retina (NSRJ and the retinal pigment epithelium (RPE). B) Intersection between eGenes identified in METR-NSR and the EyeGex retina eQTL study. For eGenes which were present in METR-NSR and EyeGex, we compared the top eQTL for each eGene to identify eQTLs which were replicated in both studies; aGenes where the top eQTL from each study was in high LD with each other (r2 > 0.8) and eGenes where the top NSR hit was novel. For a subset of eQTLs that were tested for LD, the LD score was unavailable. C) Comparison between METR-eQTLs and GTEX eQTLs indicates the number of NSR-specify:, RPE-specific, NSR & RPE-specific and non-eye specific eQTLs (left) and eGenes (right) from both tissues in our study

We compared the top METR-NSR eQTL for each eGene identified in this study with a published retina-specific eQTL dataset from the EyeGEx project (Ratnapriya *et al*., 2019) to identify: (1) eQTLs identically replicated in the METR-NSR eQTLs; (2) METR-NSR eQTLs that impacted eGenes previously described but had alternative eVariants in high linkage disequilibrium (LD) with findings from EyeGEx, and (3) previously unreported METR-NSR eQTLs, including newly identified eGenes in NSR (**Figure 2B**). Of note, our cohort excludes individuals with late-stage AMD, whereas EyeGEx includes late-stage AMD eyes. We report 5,993 NSR eGenes which were previously described by EyeGex, of which 433 eGenes share identical top eVariants and 1,700 have top eVariants in high LD with previously identified top eVariants (r^2^> 0.8). We identified 353,385 novel eQTLs in eGenes that were previously described by EyeGex and 429,377 novel eQTLs in 3,966 newly identified eGenes. Importantly, we replicate 13 eQTLs that have previously been reported to impact genes that are implicated in increased risk of AMD (***Table 1***).

**Table 1.**
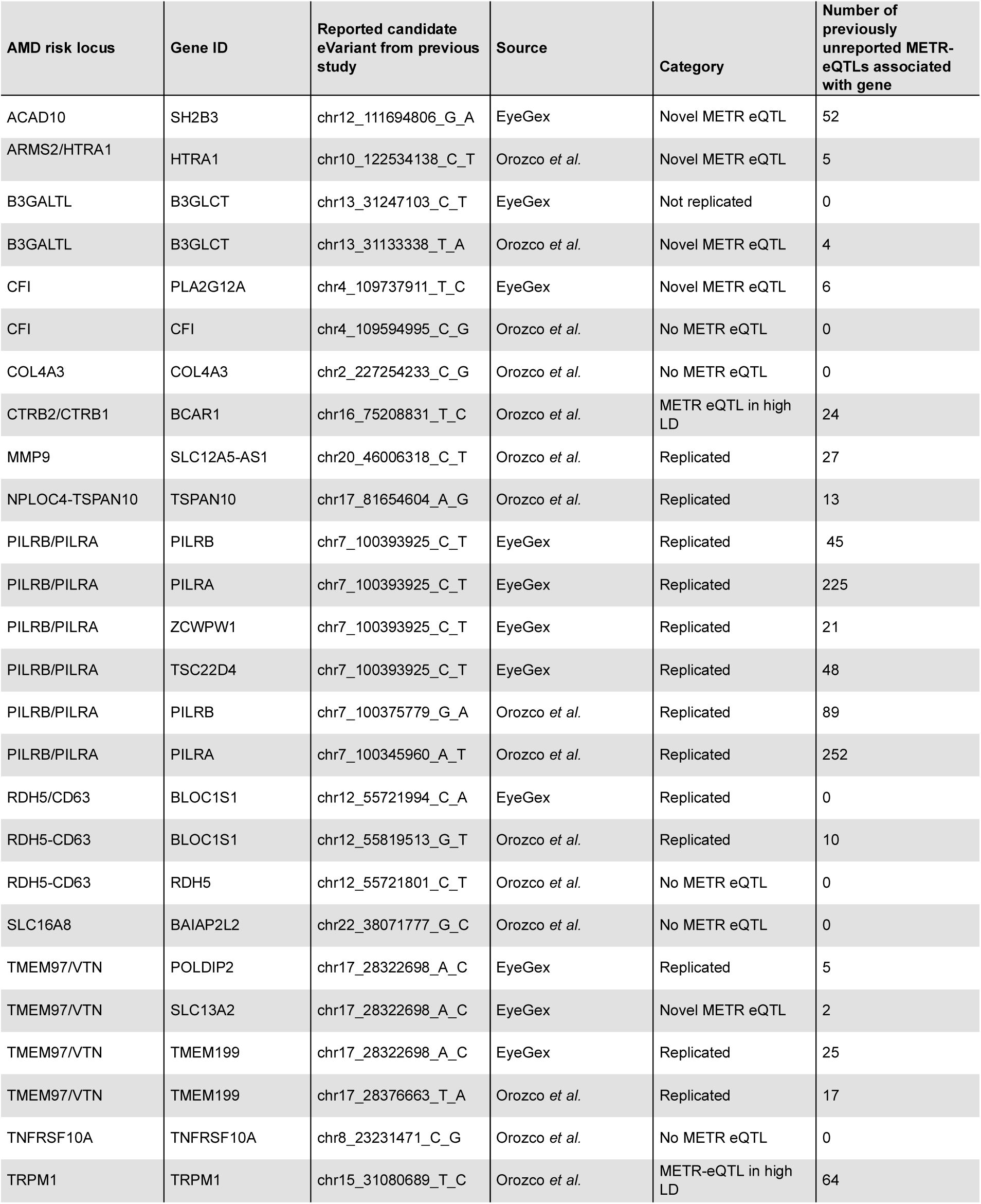
Replication of eQTLs that impact AMD risk genes and were identified as lead candidates by Ratnapriya et al. (2019) and Orozco et al. (2020) in the METR-eQTL dataset. 26 eQTLs across 14 AMD risk loci have previously been identified. Of these, 15 were replicated or in high LD (r^2^>0.8) with METR-eQTLs. Novel METR-eQTLs are identified for 5 genes previously implicated in AMD risk. Definitions utilised in table: ‘Replicated’, lead candidate variant from other studies is also identified in this study; ‘METR eQTL in high LD’, lead candidate variant from other studies is in high LD with an eQTL identified in this study; ‘Novel METR eQTL’, the lead candidate variant from other studies is not replicated in this study, additional and previously unreported eVariants are identified in this study but they are not in high LD with the previously reported lead candidate variant; ‘No METR eQTL’, the gene is not identified as an eGene in this study; ‘Not replicated’, we identify eQTLs for the gene in this study but the lead candidate variant from other studies is not replicated nor is it in high LD with an eQTL identified in this study.

### Over 800 eGenes are newly identified in the NSR and RPE

To evaluate the tissue-specificity of our dataset, we compared the METR-eQTLs with non-eye specific eQTLs from the GTEx project (**Figure 2C**). We identified 337,424 METR-eQTLs (22.7%) and 916 eGenes (8.7%) that had not been previously identified by GTEx (**Figure 2C**); 251,685 (74.6%) of these eQTLs have not been previously described as eQTLs in NSR or RPE previously. Of the novel eGenes, 479 (57.9%) encoded lncRNAs and 5 had previously been associated with a known eye-disease (*HPS4, ACO2, CRX, CRYAA, PEX26*). We evaluated the degree of similarity between METR-eQTLs and eQTLs from each GTEx tissue using the Intersection over Union (IoU) statistic, which accounts for the wide variation in the number of eQTLs from different tissues (***Supplementary figures 4 and 5***). Brain cortex had the highest level of eQTL similarity to our dataset (IoU = 0.28), and 5 of the top 10 most similar tissues were from the brain.

### Genetic variants driving expression profile differences are enriched in candidate cis-regulatory elements (cCREs), with highest enrichment in retina-specific cCREs

To understand whether eQTLs were enriched for putative regulatory regions, we compared locations of METR-eVariants to cell-type agnostic cis-candidate regulatory elements (cCRE) available through ENCODE (V3). METR-eVariants were enriched in cell-type agnostic promoters (p= 8.05×10^-19^) and proximal enhancers (p = 8.48×10^-26^), compared to control variants matched for allele frequency and gene density. There was no enrichment of eVariants in distal enhancers, CTCF binding sites or DNase-H3K4me3 sites (**Figure 3A**).

**Figure 3.**
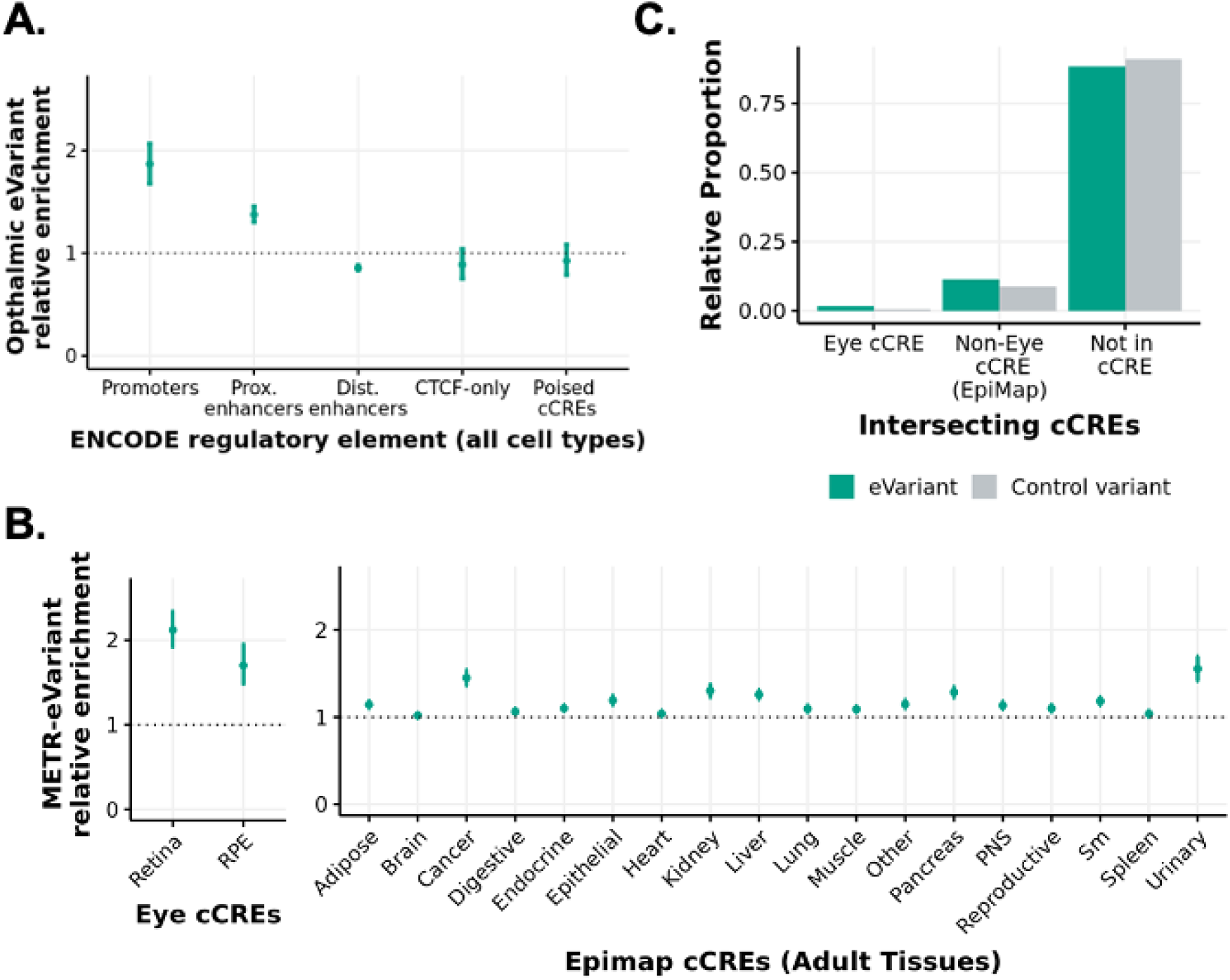
Enrichment of eye eVariants in previously annotated candidate cis-regulatory elements (cCREs) **A)** Bootstrapped relative enrichment of aye eVariants which intersect with cCREs from the ENCODE cCRE registry (VJ) compared to control variants. Relative enrichment is defined as the ratio of eVariants to control variants which intersect with each element. tn each bootstrapping iteration, random subsets of eVariants and control variants (subsets of non-eVariant matched for gnomad AF and gene density) were intersected with each cCRE group.. E.rror bars indicate the 2.5-97.5% confidence intervals. **B)** Rel’ative enrichment of eye eVariants which intersect with cCREs from retina and RPE (Cherry et al., 2020) and cCREs from adult tissues in EpiMap compared to control variants matched for gnomad AF and gene density. Error bars indicate the 2.5-97.5% bootstrapped confidence intervals. **CJ** Relative propomon of eVariants and non-eVarlants in the NSR and/or RPE which intersect with annotated retina cCREs (Cherry et al., 2020), non-retina oCREs (EpiMap) and neither.. Control variants refer to those which were included in eQTL mapping (MAF > 2.5% and AC > 10) but were not associated with any eQTLs in NSR or RPE.

When stratified by cell-specific regulatory regions, bootstrapping analysis indicated a significant enrichment of METR-eVariants in NSR-specific (p-value = 4.52×10^-28^) and RPE-specific cCREs (p-value = 8.74×10^-10^) compared to control variants matched for allele frequency and gene density (number of gene TSSs within 1Mb of variant). Non-eye cCREs from adult tissues in EpiMap were also enriched for METR-eVariants, although the enrichment was lower than in the NSR and RPE (**Figure 3B**). Despite the relative enrichment in annotated regulatory loci, most METR-eVariants (88.2%) do not overlap with any previously characterised cCREs (**Figure 3C**).

### Properties of METR-eQTLs differ between known disease-related genes and non-disease-related genes

230 METR-eGenes have a known association with eye disease in the EyeG2P resource (Lenassi *et al*., 2023). We compared properties of disease-related eGenes in EyeG2P (‘eye disease eGenes’) with all other METR-eGenes (‘eye non-disease eGenes’). We observed significantly lower expression variability across samples for eye disease eGenes compared to eye non-disease eGenes (*p*<2.2×10^-16^) (**Figure 4A**). Eye disease eGenes were associated with significantly fewer eQTLs per gene (*p*=7.8×10^-3^) (**Figure 4B**). Additionally, eQTLs associated with eye disease eGenes have a significantly lower impact on gene expression (*p*<2.2×10^-16^) (**Figure 4C**), and significantly higher allele frequency (AF, gnomAD v4) compared to eye non-disease eVariants (*p*<2.2×10^-16^) (**Figure 4D**). For both eye disease eGenes and eye non-disease eGenes, there is a negative correlation (*p*<2.2×10^-16^) between eVariant allele frequency and the impact of each eQTL on gene expression (**Figure 4E**). These findings are consistent with the hypothesis that eVariants which are more common in the population have lower effect sizes on gene expression compared to rarer eVariants (min eVariant allele frequency = 2.5%), and are suggestive of a selective bias against rarer eVariants impacting known eye disease genes.

**Figure 4.**
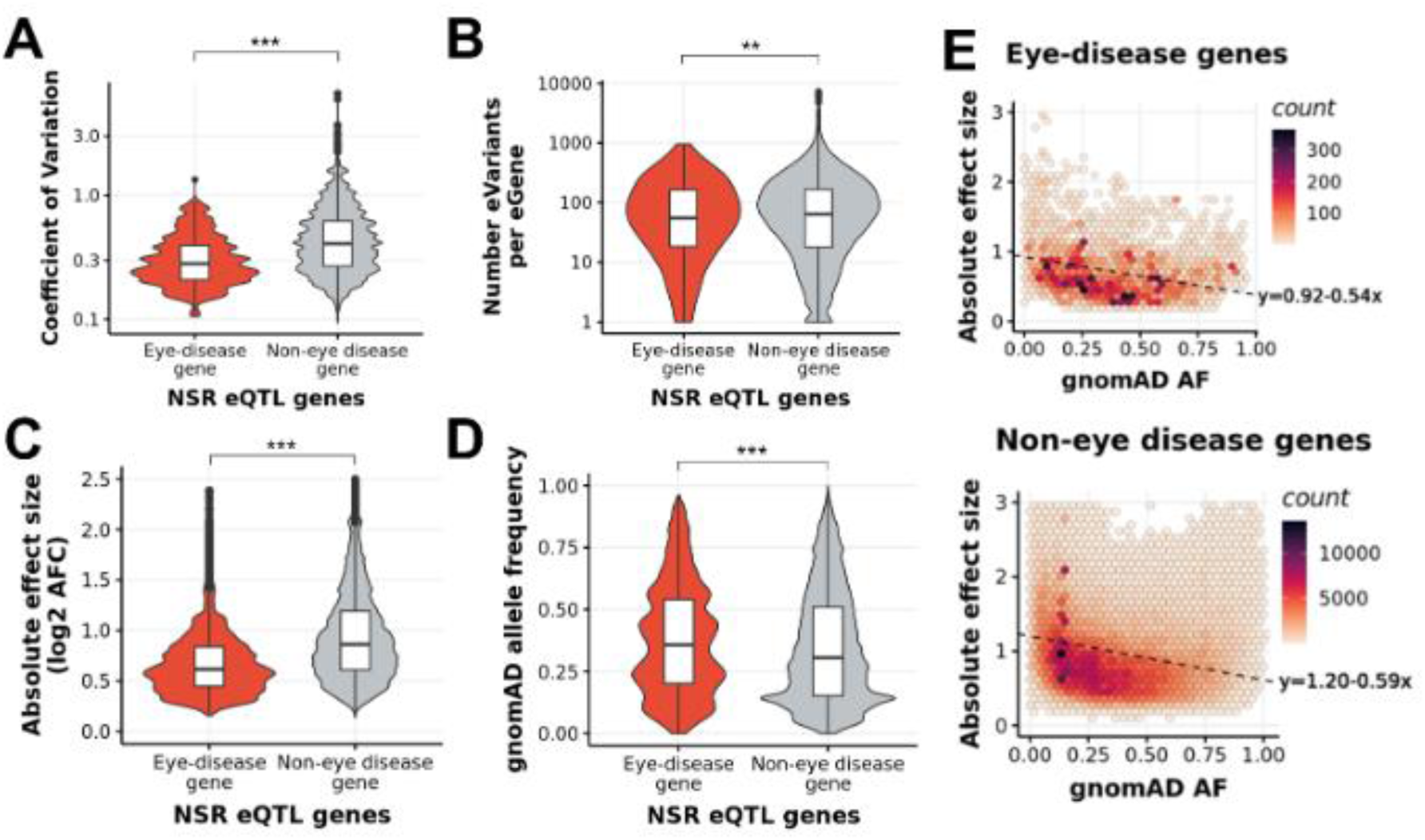
Differences, In NSR eQTLs associated with known eye disease genes and’ genes whlc.h are not attributed to eye diseases (non--eye disease genes) **A)** Genes associated with eye diseases on EyeG2P have lower coefficients of variation than non-eye disease genes which indicates lower expression variability across samples. **B)** On average, known eye disease genes have fewer eVariants per gene than non-eye disease genes. CJ. The impact on gene expression (measured in absolute Jog2 allelic fold change) of each eQTL associated with a known eye disease gene is lower than the impact of eQTLs associated with non-eye disease genes. **D)** The allele frequency on gnomAD for the eVariants associated with eye disease genes is higher than non-eye disease genes **E)** There is a negative linear relationship between the allele frequency of an evariant, and the impact on gene expression of the associated eGene (measured in Jog2(allelic fold change)) for eQTLs associated with eye disease genes (top)’ and non-eye disease genes (bottom)

### Rare variants are plausible drivers of transcriptomic outliers in NSR and RPE

We utilised the DROP workflow (Yépez *et al*., 2021) to identify statistical outlier events within the METR transcriptome datasets, including expression, splicing and allelic imbalance outliers (***Table 2***). We identified 1,051 unique METR expression outliers (METR-eOutliers) (adjusted *p*<0.05); 728 of these events were in the NSR, 443 in the RPE, and 120 eOutliers were found in both tissues.

**Table 2.** Transcriptome outliers detected in the METR-cohort. The DROP pipeline was utilised to identify three types of transcriptome outliers (expression, splicing and monoallelic expression) in the NSR and RPE.

| Type of outlier | Expression |  |  | Splicing |  |  | Allelic balance |  |  |
| --- | --- | --- | --- | --- | --- | --- | --- | --- | --- |
|  | NSR | RPE | Shared | NSR | RPE | Shared | NSR | RPE | Shared |
| Unique events (count) | 728 | 443 | 120 | 8,610 | 8,923 | 469 | 37,230 | 106,116 | 9,888 |
| Unique genes (count) | 702 | 439 | 137 | 6,095 | 6,270 | 2,972 | 7,893 | 17,167 | 7,410 |
| Median number of impacted genes per sample | 3 | 1 | 0 | 12 | 6 | 1 | 173 | 138 | 53.5 |
| IQR (Q1, Q2) | (1, 4) | (0, 2) | (0, 1) | (8, 18) | (3, 24) | (0, 2) | (146, 203) | (71, 1,003) | (34, 87) |
| Range (min, max) | (0, 64) | (0, 46) | (0, 24) | (2, 1,948) | (0, 1,258) | (0, 209) | (102, 2,823) | (14, 4,308) | (8, 214) |

For each eOutlier, we were able to harness paired genomic data to identify candidate rare variants potentially driving aberrant expression profiles. We leveraged a hierarchical framework and a probabilistic model to prioritise candidate rare genetic variants driving changes in expression. This identified 230 (23%) eOutlier events likely driven by protein-coding variants and 314 (31%) events with non-coding candidate variants (**Supplementary table 5**).

### Rare variants predicted to have a functional impact are identified for 50% of eOutlier events in NSR and RPE

First, we applied a hierarchical framework to identify rare SVs, CNVs and SNVs which were predicted to result in loss-of-function (pLoF, including frameshift, nonsense and start/stop site loss variants) or were expected to disrupt a nearby non-coding regulatory region (**Supplementary figure 6**). Following this approach, we identified candidate functional variants driving 528 eOutlier events (50.2% of all METR-eOutliers) (**Figure 5A; *Supplementary table 5***). Of these, 131 METR-eOutliers were co-occurring with a SV or CNV impacting the coding-sequence of the outlier gene (77 NSR-only,12 RPE-only and 42 in both tissues), and 98 with a pLoF SNV (77 NSR-only, 6 RPE-only and 15 in both tissues) impacting the same gene. For those eOutliers not explained by an SV, CNV or pLoF SNV disrupting the coding sequence, we identified genetic variants in 71 eOutliers that were within 10Kb of the gene body and impacted an eye-specific cCRE (Cherry *et al*., 2020), including SV/CNVs (n=2) and SNVs that are rare (<0.01 AF) or absent in gnomAD (n=69). We also identified non-eye specific cCREs from EpiMap (Boix *et al*., 2021) which were disrupted by SV/CNVs (n=23) or rare SNVs (n=121) within 10 Kb of the eOutlier gene. Examples of rare variants identified through this analysis strategy are included in ***Figures 6*&7**.

**Figure 5.**
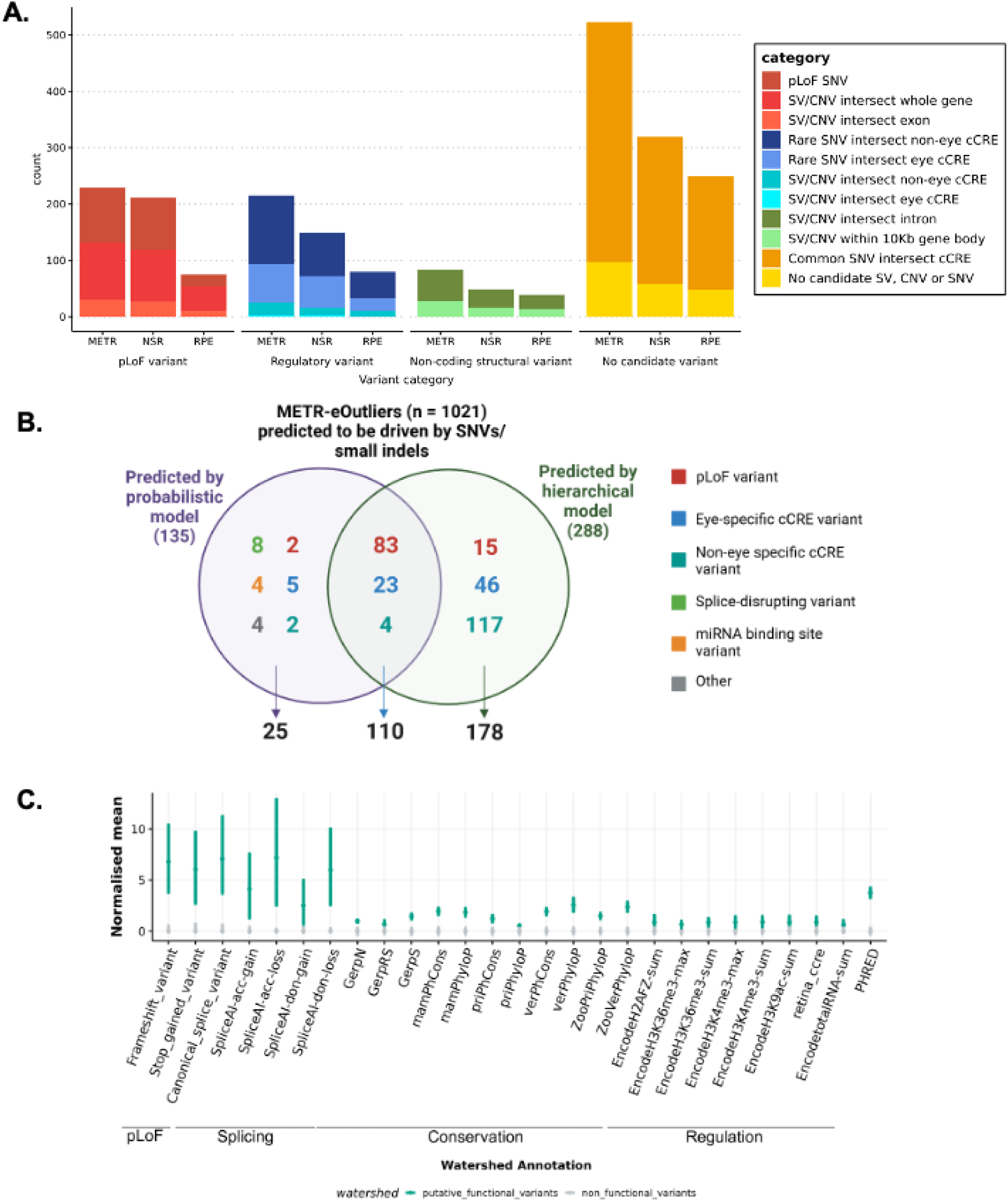
Identification of rare variants driving METR transcriptomic outliers in neurosensory retina (NSR) and retinal pigment epithelium (RPE). **A)** The hierarchical world/ow to identify candidate variants driving outlier expression identified putative functional variants driving 563 eOutlier events in NSR, RPI= or both (METR) **B)** The Watershed probabilistic model had high concordance with the hierarchical framework for the prioritization of SNVs and small indels driving eOutliers in NSR and RPE CJ Bootstrapping analysis indicates that variants which were predicted to be driving eOutliers by Watershed were enriched for pLoF variants, splicing variants, variants within regulatory elements (including retina cCRl=s) and those with high conservation scores compared to those which were not predicted to have a functional impact. Bars indicate the 95% CI.

**Figure 6.**
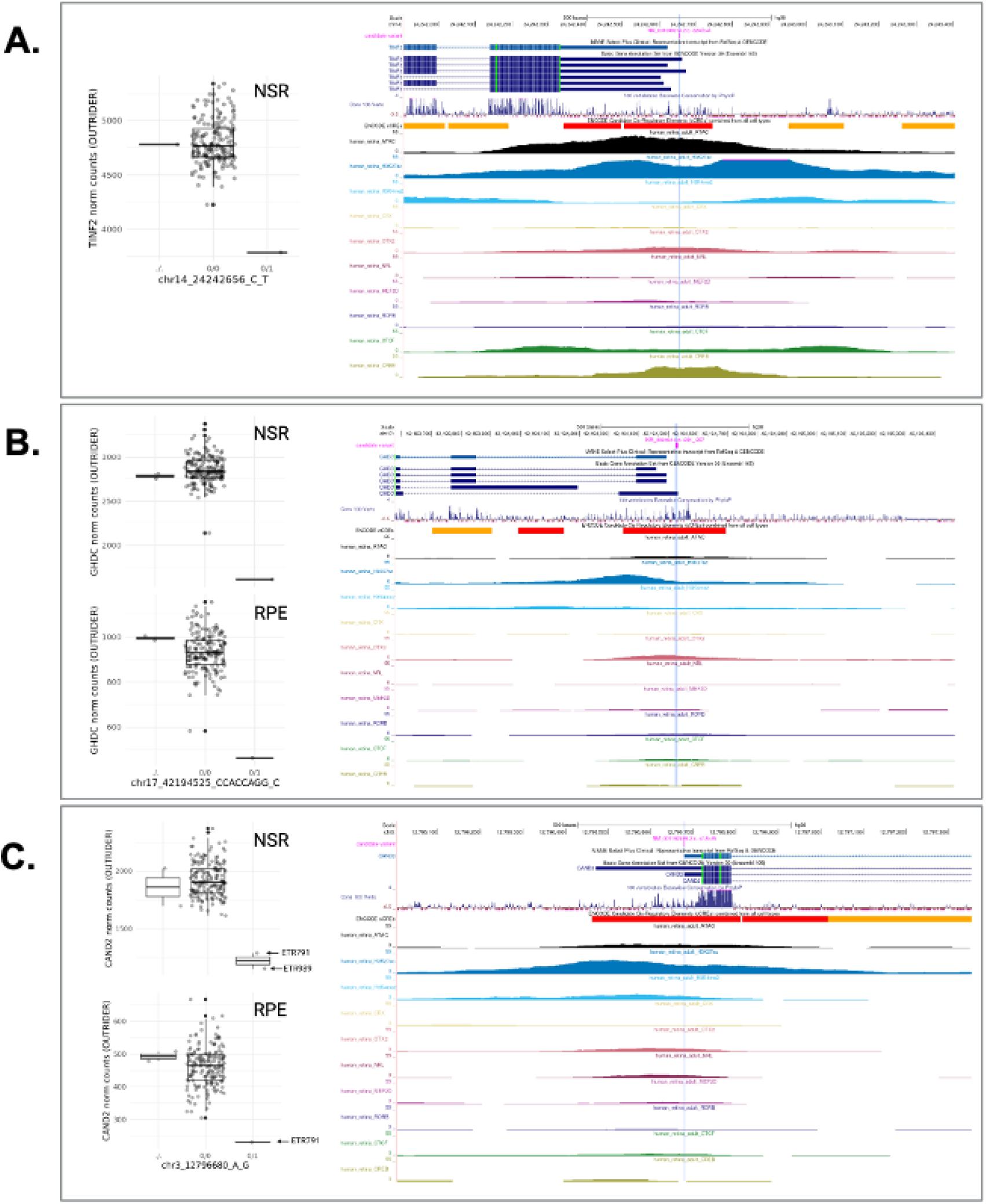
Candidate rare small variants driving METR transcriptomic outliers in neurosensory retina (NSR) and retinal pigment epithelium (RPE). In each caption, the tissue and relative outlier expression profiles calculated through OUTRIDER for individuals with missing (./.), homozygous reference (0/0) and heterozygous ultimate (0/1) genotypes are shown, alongside genome tracks displaying transcript isoforms, evolutionary conservation and candidate cis­ regulatory elements (cCREs) identified in EpiMap and Cherry et al. Box and whiskers plots show median values and interquartile ranges, with grey dots indicating normalized count values for single samples, and statistical outliers for each genotype indicated with black dots. In all examples, the variant intersects epigenomic peaks in retina and other tissues indicative of a promoter region for the MANE transcript: **A)** NM_001099274.2:c.-324G>A, TINF2; **BJ** NM_032484.4:·c.-231_237de1, GHDC, outlier expression in both NSR and RPE; CJ NM_001162499.2:c.-41A>G, CAND2, two donors carrying this heterozygous variant have outlier expression, with one donor showing outlier expression in both NSR and RPE.

### A probabilistic model demonstrates high concordance for candidate SNVs driving expression outlier profiles in NSR and RPE

Next, we applied Watershed (Ferraro *et al*., 2020), a probabilistic model that was re-trained on 6 tissue-specific outlier *p*-values from DROP. This was used in the METR transcriptome datasets to obtain posterior probabilities for SNVs and small indels that may be driving outlier expression profiles. Eye-specific cCREs were added as annotation features for Watershed (***Supplementary Table 6***), and identified 135 (13%) METR-eOutliers that were likely to be caused by nearby rare variants (posterior probability > 0.8), of which 110 (81%) were also predicted to be driven by the same variants by the hierarchical model and 11 were predicted to be driven by SV/CNVs that are not considered by Watershed (**Figure 5B**). We used bootstrapping analysis to compare the annotations associated with these variants to other rare variants which overlapped with eOutlier genes but were not predicted by Watershed to have a functional impact, observing an enrichment of canonical splice variants, frameshift variants, stop gain variants and variants predicted to disrupt splicing (**Figure 5C**). In support of other analyses described in this study, there was an enrichment of rare variants which overlap with retina cCREs, and a slight enrichment of rare variants which overlap with epigenomic marks associated with non-eye specific regulatory elements from ENCODE. In total, there were 34 eOutliers where the functional variants prioritised by Watershed intersected with a known candidate cis-regulatory region (cCRE); 28 of these cCREs were active in the eye.

## Discussion

We present a unique resource to interrogate the impact of both common and rare genetic variation on gene regulation in the human NSR and RPE. We characterised novel eQTL associations that are tissue-specific (**Figure 2**) and are enriched to known promoters and proximal enhancers (**Figure 3**). We show that eQTLs impacting genes known as a cause of genetic eye disease have different properties when compared to those genes which are not known as a cause of eye disease (***Figure 4***). We also identify candidate non-coding rare variants, SVs and CNVs which impact cCREs and represent plausible drivers of outlier expression profiles in human NSR and RPE (***Figure 5***). The METR resource can be used alongside other multi-omic datasets to facilitate discovery of novel eye-specific regulatory elements, including those implicated in common (e.g. AMD) and rare (e.g. IRDs) genetic disorders impacting the retina.

The cohort of 201 human donors described in this study represents the first dataset, to our knowledge, to pair whole genome sequencing with high-depth RNA sequencing data from the NSR and RPE. Previous studies have developed lower-depth RNA sequencing from the NSR alongside genotyping arrays (Ratnapriya *et al*., 2019; Orozco *et al*., 2020; Strunz *et al*., 2020) and this has enabled the characterisation of eQTLs in the retina including preliminary data supporting the role of a limited number of eQTLs in AMD (***Table 1***). The RNA and whole genome sequencing datasets developed for this study are from a cohort of individuals without clear signs of late-stage AMD, and enable novel biological insights, beyond those described previously.

Firstly, we were able to assess whether previously characterised eQTLs are replicated utilising alternative methods and technologies in an independent cohort of individuals without late-stage AMD. As gene expression profiles have been shown to be significantly disrupted during AMD pathogenesis (Voigt *et al*., 2022; Orozco *et al*., 2023), it is important to identify eQTL signals that are amplified or disrupted by broader changes in transcriptome profiles associated with AMD, as well as those that remain consistent within a cohort of individuals without clear signs of late-stage AMD. Overall, we show high levels of replication of eQTL findings from Ratnapriya et al, with 5,993 identical eGenes and replication of 13 eQTLs previously implicated with a role in AMD (***Table 1***), including *PILRB* which has recently been shown to lead to photoreceptor dysfunction in mice when function is impaired (Dey *et al*., 2025). Notably, we identified 5 novel QTLs for genes previously implicated in AMD (*ACAD10, HTRA1, B3GLCT, PLA2G12A, BAIAP2L2*) and 4 genes implicated in AMD without replication of a previously characterised eQTL (*CFI, COL4A3, RDH5, TNFRSF10A*). This suggests that differences in the approaches undertaken and/or cohort composition, e.g. AMD status, cohort size and/or genetic ancestry, impacts the influence of genomic drivers on expression of these genes.

Second, variants which are rare in the population or unique to individuals have been demonstrated to drive drastic changes in expression profiles, so called ‘expression outliers’, across different tissues (Li *et al*., 2017; Ferraro *et al*., 2020). The use of complete genomic sequencing in this cohort, achieving a median coverage of 36x, has enabled the characterisation of a greater diversity of genetic variation than has previously been studied in the context of expression drivers in the NSR and RPE, and identified thousands of new regions which can be interrogated for rare variation within disease cohorts (Ellingford *et al*., 2022). Using two distinct variant prioritisation approaches, we describe rare variants in the general population, including SVs, CNVs and small variants that are the most likely drivers of expression outliers in these tissues (***Figure 5***). Whilst functional follow-up is required to confirm the disruptive role of these variants, we identify 578 variants that may be causative of pronounced changes in expression profiles in the human retina, accounting for over 50% of observed outlier events. This includes 272 rare variants predicted to cause loss-of-function and 299 that intersect with non-coding regions, providing further insight into the regions regulating gene expression in the human retina (Cherry *et al*., 2020), including examples of small variants presented in ***Figure 6*** and SV/CNVs presented in ***Figure 7***. Other recent studies have identified outlier-associated non-coding rare variants that contribute to common disease predisposition (Smail *et al*., 2022) and underpin rare genetic disorders (Wakeling *et al*., 2022; Tenney *et al*., 2023). Moreover, non-coding variation has been identified as a cause of genetic ophthalmic disorders in untranslated regions (Dueñas Rey *et al*., 2024), retina-specific exons (Vig *et al*., 2020), promoters (Daich Varela *et al*., 2023), distal enhancers (Small *et al*., 2016) and non-coding genes (Quinodoz *et al*., 2025) expressed in the NSR and RPE. With the increasing availability of genomic sequencing datasets for the diagnosis and discovery of genetic disorders (Turnbull *et al*., 2018; The 100,000 Genomes Project Pilot Investigators, 2021), including ophthalmic conditions (Ellingford *et al*., 2016), the data developed in this study is timely and provides an opportunity, alongside other complementary datasets (D’haene *et al*., 2024), to identify new pathogenic mechanisms underpinning genetic disorders.

**Figure 7.**
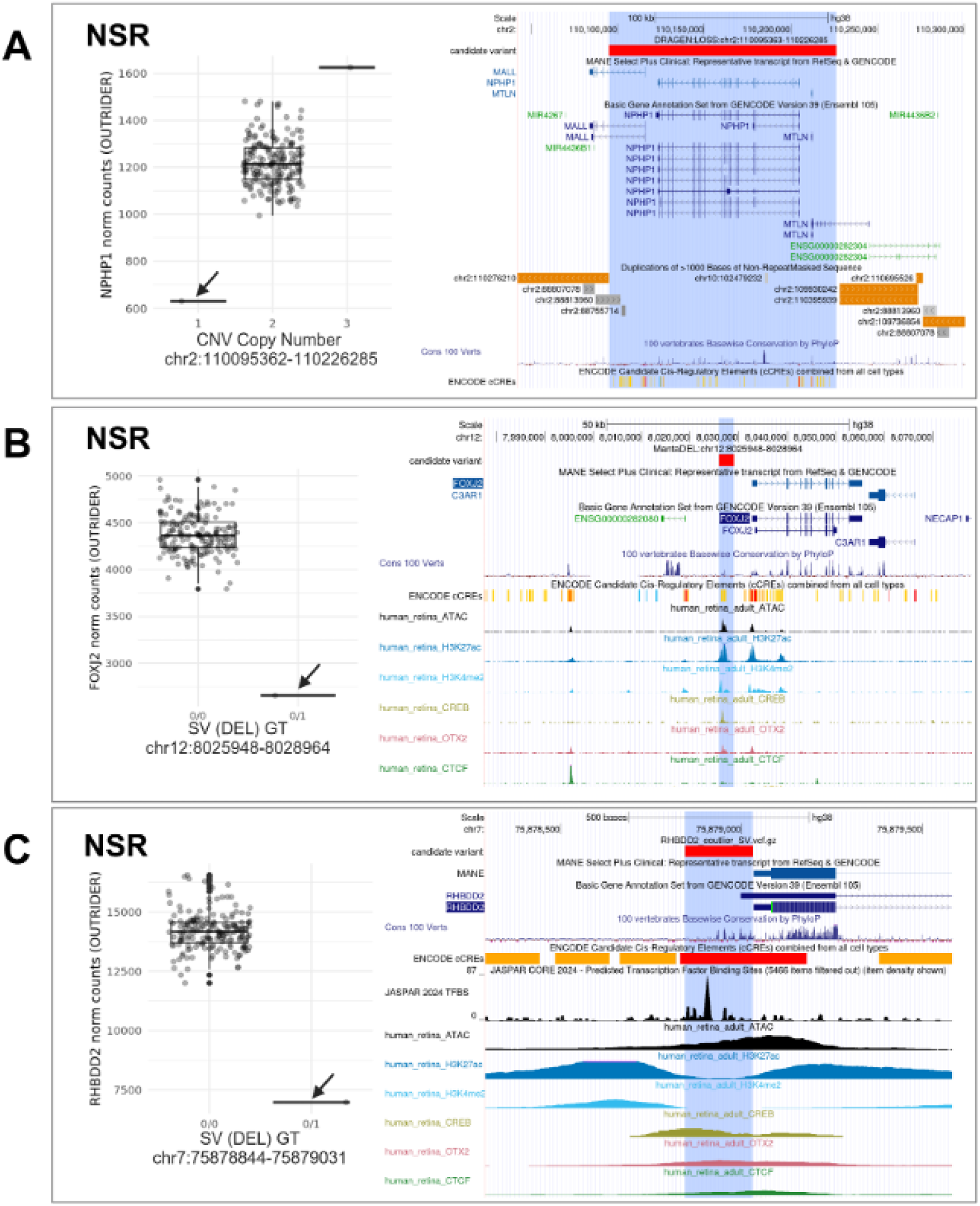
Candidate structural and copy number variants driving METR transcriptomics outliers In neurosensory retina (NSR).. Jn each caption, the tissue and relative outlier expression profiles calculated through OUTRIDER for individuals are shown, with: **(A)** copy number states 1, 2 and 3; and **(B,**C) homozygous reference (0/0) and heterozygous alternate (0/1) genotypes. Genome tracks displaying transcript isoforms, evolutionary conservation and candidate cis­ regulatory elements (cCREs) identified in EpiMap and Cherry et al are included alongside: **(A)** segmental duplication regions; and **(B,C)** JASPAR transcription binding sites. Box and whiskers plots show median values and interquartile ranges, with grey dots indicating nomialised count values for single samples, and statistical outliers for each genotype indicated with black dots. In **(A)** CNIVs encompassing1a known eye disease gene (NPHP1) are shown to cause drastic changes in expression in NSR. In **(B)** and **(C)** deletion, s are shown to impact cCREs proximal lo the transcription start sites of genes expressed in NSR: (IB) 3Kb deletion −3.5Kb upstream of FOXJ2; (C) 187bp deletion impacting1minimal promoter reg1ion of RHBDD2..

Third, we have generated high-coverage RNAseq datasets achieving, on average, 139 million uniquely mapping reads for NSR and 62 million uniquely mapping reads for RPE. Previous studies have developed lower coverage RNAseq datasets, for example, EyeGEx (Ratnapriya *et al*., 2019), Orozco *et al* (Orozco *et al*., 2020) and Pinelli *et al* (Pinelli *et al*., 2016) generated 33, 30 and 72 million sequencing reads per sample, respectively. Previous studies have remarked on the level of transcript diversity in NSR (Farkas *et al*., 2013), and highlighted the advantage of high-depth RNAseq in this context. In comparison to EyeGEx, our high coverage approach elevates the number of observable protein-coding genes by 23% (from 13,662 to 16,765) and newly identifies 3,663 eGenes. This enables observation of patterns in gene expression at increased resolution and has granted insight into the trends associated with genes previously implicated in genetic disorders impacting vision. Overall, we observed that eGenes that have been characterised as a cause of genetic eye disease (Lenassi *et al*., 2023) have lower expression variability across individuals than non-disease genes (***Figure 4***), suggesting that regulation of these genes is more tightly controlled in NSR and RPE. The role of eQTLs in genetic disorders remains incompletely understood. For example, whilst some studies have shown eQTLs contribute to onset, penetrance and expressivity (Castel *et al*., 2018; Einson *et al*., 2023), including genetic disorders impacting the eye (Michaud *et al*., 2022), others have found limited evidence for their role in neuronal genetic disorders (Rio Frio *et al*., 2008; Wigdor *et al*., 2024). Here, we observe that eQTL variants which were associated with changes in expression of eye-disease genes had significantly lower effect sizes and their allele frequency was higher than eQTLs impacting genes that have not previously been implicated in eye disease. Intuitively, the absence of rarer and higher impact eVariants amongst a population of individuals without signs of genetic eye disease suggests constraint on genetic variation with these properties.

Finally, as our cohort includes 158 individuals with RNA extracted and sequenced from both NSR and RPE datasets, this enables further insights into the expression patterns and regulatory architecture of these tissues, unbiased by sample preparation methods and/or differences between individuals, e.g. genomic background. Our data newly identifies 916 eGenes in NSR and RPE compared to those characterised in other tissues (THE GTEX CONSORTIUM, 2020). Whilst over 20,000 genes are differentially expressed between NSR and RPE samples in our cohort, we observe high level of overlap, including 86% of RPE eGenes and 32% of NSR eGenes. These data further support the high level of overlap previously observed for active enhancers and promoters between RPE/choroid and NSR (Cherry *et al*., 2020).

Whilst the findings of this study have enhanced our understanding of genomic regulation in human NSR and RPE, other approaches that utilise single-cell (Lukowski *et al*., 2019; Menon *et al*., 2019; Orozco *et al*., 2020; Yan *et al*., 2020; van Zyl *et al*., 2022) single-nuclei (Liang *et al*., 2019; Monavarfeshani *et al*., 2023) and spatial (Choi *et al*., 2023; Dorgau *et al*., 2024) transcriptomic approaches enable increased precision to understand gene expression in specialised retinal layers and cell types. These approaches are particularly advantageous for the NSR, which is a highly heterogeneous tissue comprised of several specialised layers and neuronal cell types, including photoreceptors, bipolar cells, amacrine cells and horizontal cells (Masland, 2012). To overcome potential shortcomings of the bulk RNAseq approach adopted in this study, we performed deconvolution analyses to estimate the relative sample composition against single-nuclei RNA-sequencing (Monavarfeshani *et al*., 2023). Given the complexity associated with retinal tissue dissection and storage (McHarg *et al*., 2015; Cabral *et al*., 2017), the deconvolution approach also enabled confirmation of tissue sample integrity alongside differential expression profiles (***Figure 1C***). Bulk RNAseq from NSR had representation, as expected, from diverse cell types with significant enrichment towards rod photoreceptors and retinal astrocytes, representing >50% of the estimated cellular make-up of most samples. In keeping with current understanding of retinal ageing (Gao and Hollyfield, 1992) there is observed significant loss of rod photoreceptors with age (***Supplementary Figure 2B***). However, deconvolution is naturally limited by the relative differential transcriptional activity between cell types, and is complicated by cell types with similar transcription profiles, for example, between Müller glia and retinal astrocytes (Yan *et al*., 2020). Moreover, the retina is known to have cyclic patterns of gene expression, related to both circadian rhythm and natural function, i.e. response to light (Bhoi *et al*., 2023), and as such there is incomplete molecular understanding of all cell types present in the human retina (Monavarfeshani *et al*., 2023). Overall, these data support the integrity of the RNAseq dataset developed in this study and confirms datasets are representative of major cell types in the retina.

Taken together, the data presented in this study provide new insights into the genomic control of gene regulation in the human retina. We build upon previous understanding through replication of previous eQTLs in a cohort of individuals without clear signs of late-stage AMD, characterise hundreds of new genes under genomic regulation, and provide insights into the role of rare variants, SVs and CNVs in disruption of gene expression in these specialised tissues that enable vision. Future studies utilising this resource will continue to develop understanding of the expression profiles and the role of non-coding genetic variation in the onset and presentation of genetic disorders impacting vision.

## Supplementary Figures

**Supplementary figure 1.**
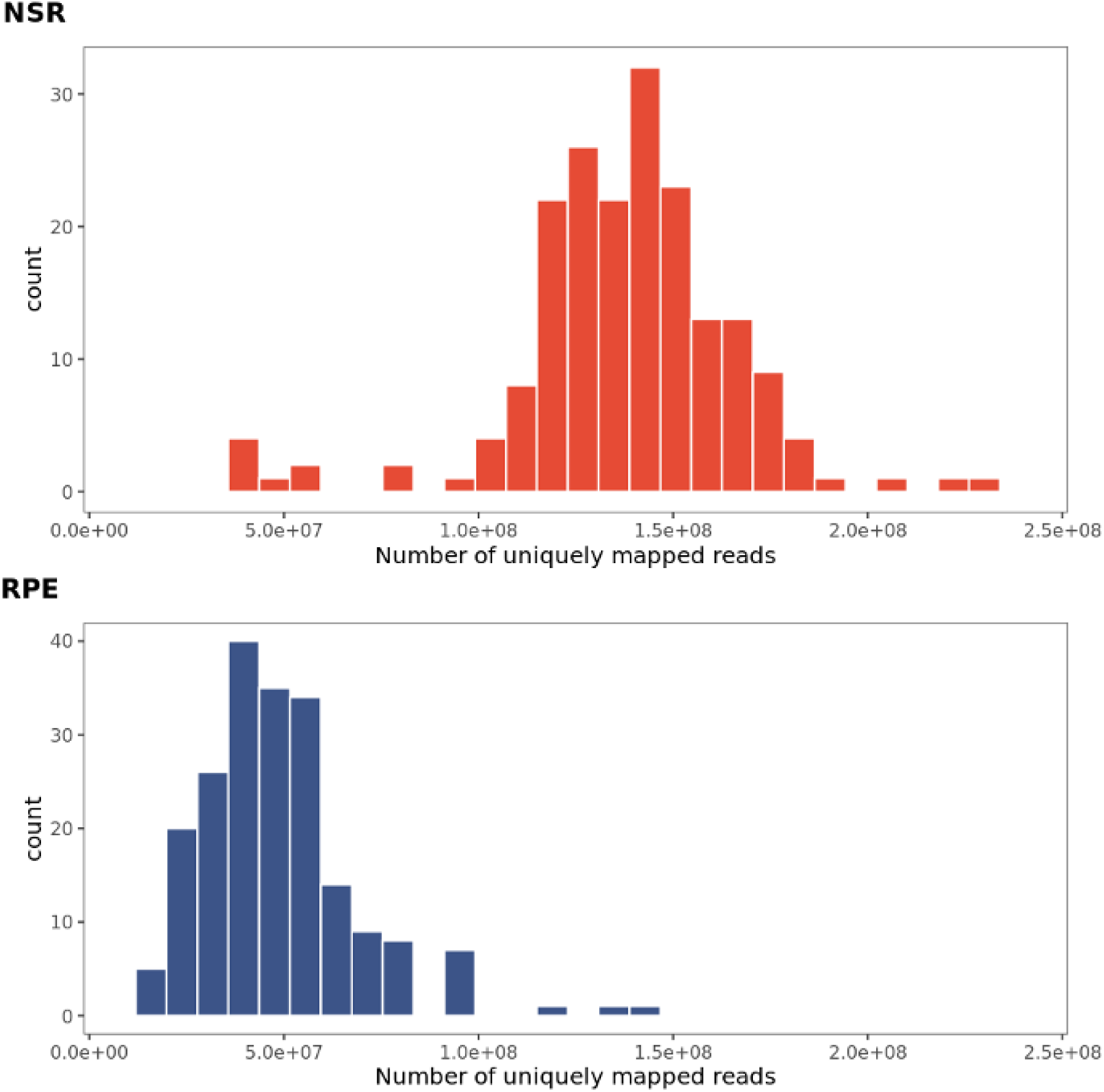
Number of uniquely mapped reads in NSR (n = 183) and RPE (n = 116) samples from the METR-GT cohort

**Supplementary figure 2.**
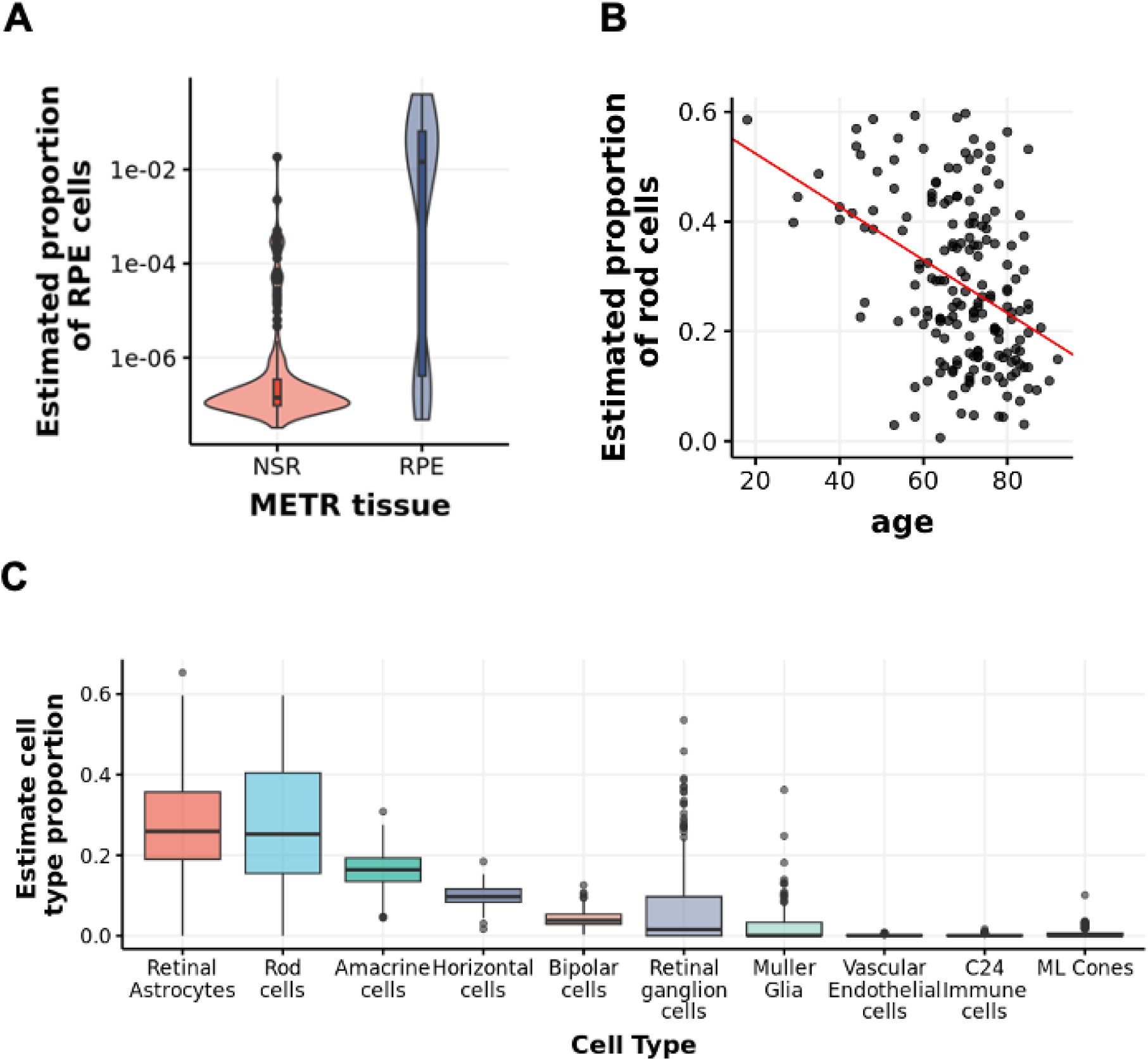
Cell type deconvolution of METR bulk RNASeq samples A The estimated proportion of RPE cells in METR-RPE samples is higher than in the METR-NSR cohort. B. The estimated proportion of rod cells decreases in samples from older donors. C. The estimated proportion of individual cell types across the METR-NSR cohort.

**Supplementary figure 3.**
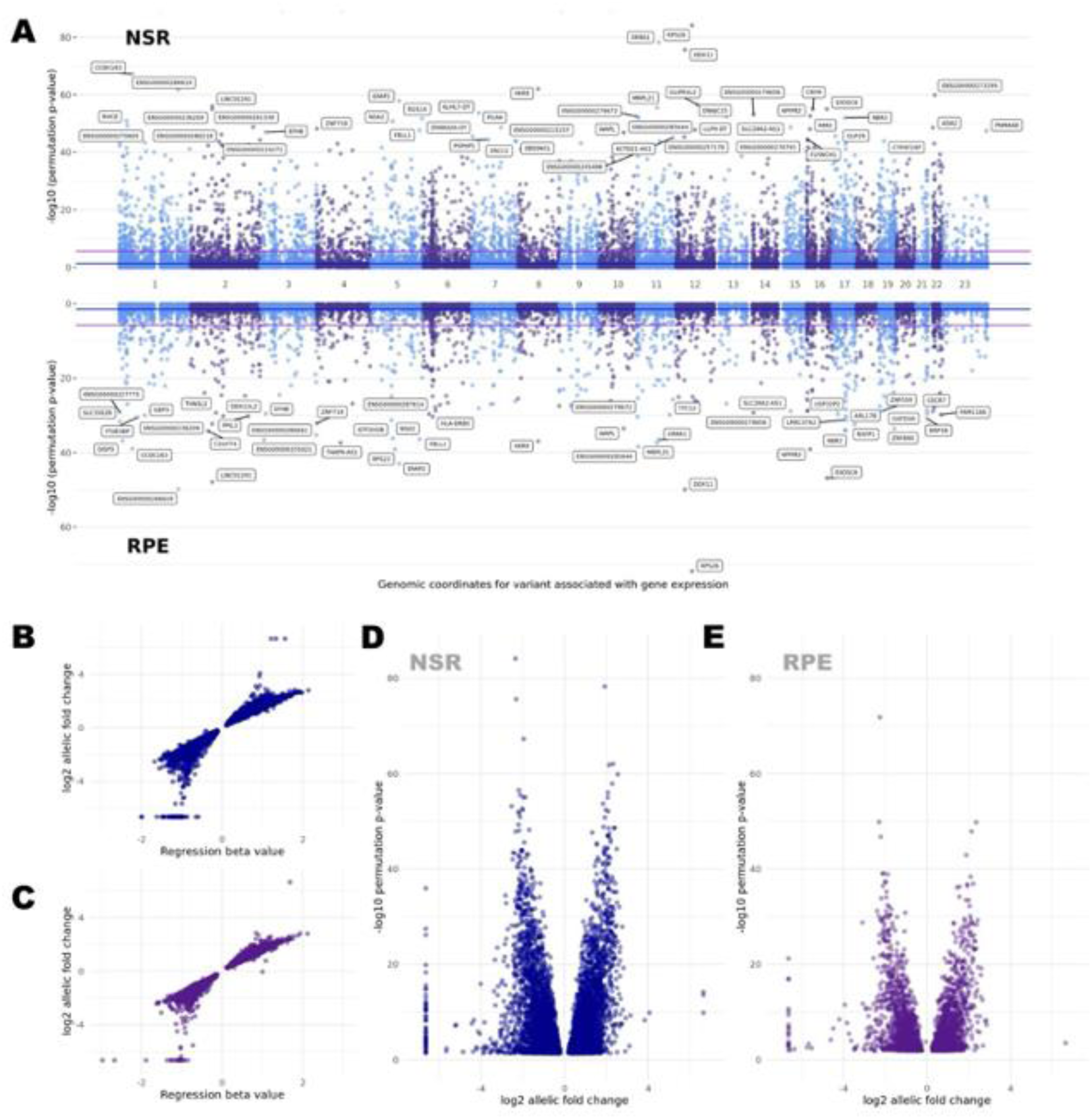
Cis-eQTL mapping in NSR and RPE based on a permutation model to identify the top eQTL candidate per gene. **A)** Manhattan plot of the top cis-eQTLs per gene in the NSR (top) and the RPE (bottom). The blue line indicates the False Discovery Rate (FDR) threshold of 5% used to identify genes with a significant (aGenes). In the NSR we identified B,609 eGenes and 3,229 in the RPE. The purple line indicates the more conservative Bonferroni corrected p-value threshold 0.05). **B)** Relationship between the regression beta values computed for each of the top eQTLs in the NSR and the Jog2 allelic fold change. The magnitude of the beta value has no direct biological interpretation as it is computed based on transformed expression values. **C)** Relationship between the regression beta values computed for each of the top eQTLs in the RPE and the log2 allelic fold change. **D** Volcano plot of the log2 allelic fold change for each of the top eQTLs in NSR **E)** Volcano plot of the log2 allelic fold change for each of the top eQTLs in RPE

**Supplementary figure 4.**
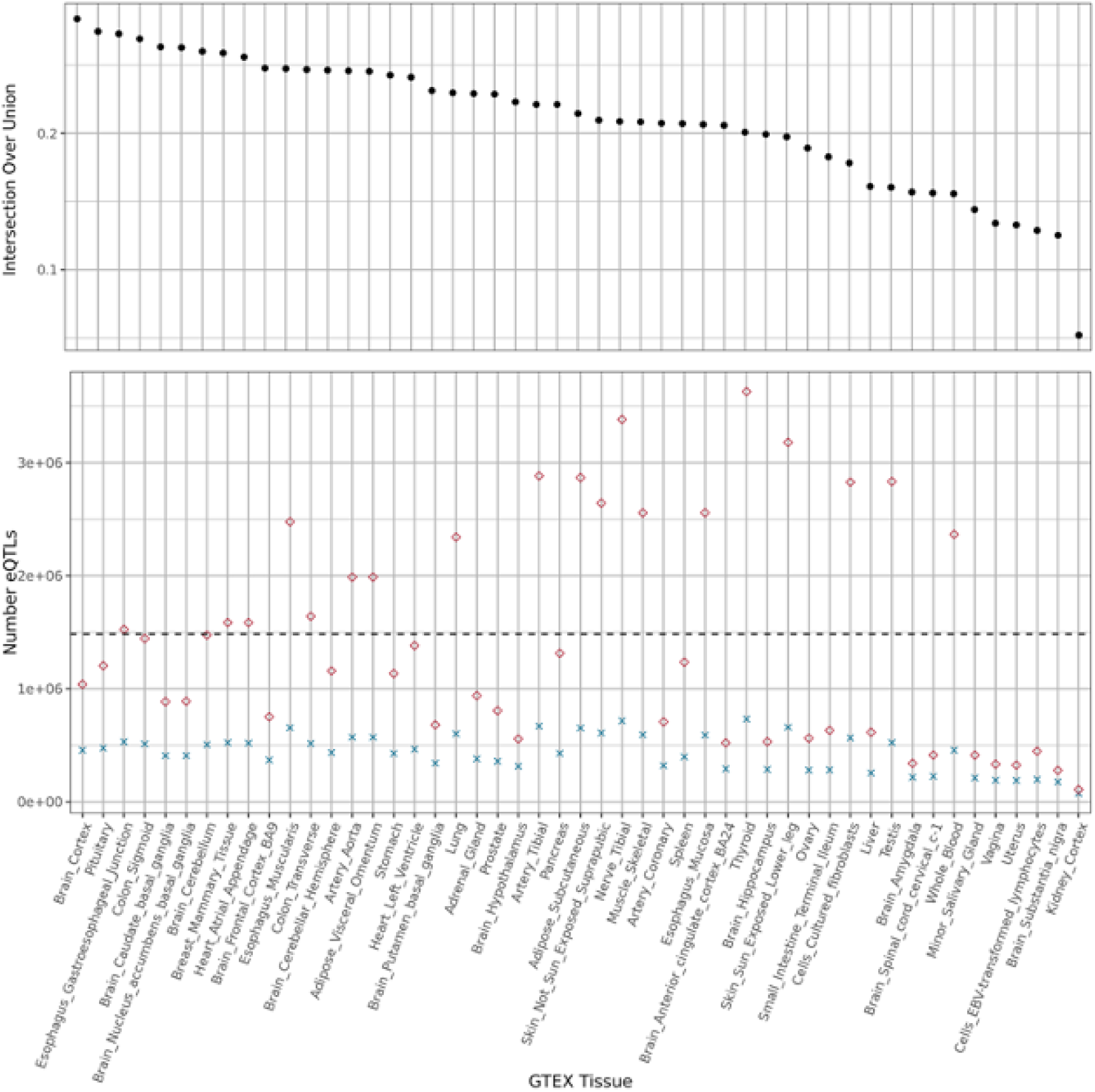
Intersection of the eQTLs identified in NSR and/or RPE with each GTEx study (vB). The bottom plot indicates the number of shared eQTLs in NSRIRPE and each tissue (cross) and the overall number of eQTLs identified in each GTEx study (circle). The top plot indicates relative proportion of eQTLs identified in each lassie and our study (intersection over union).

**Supplementary figure 5.**
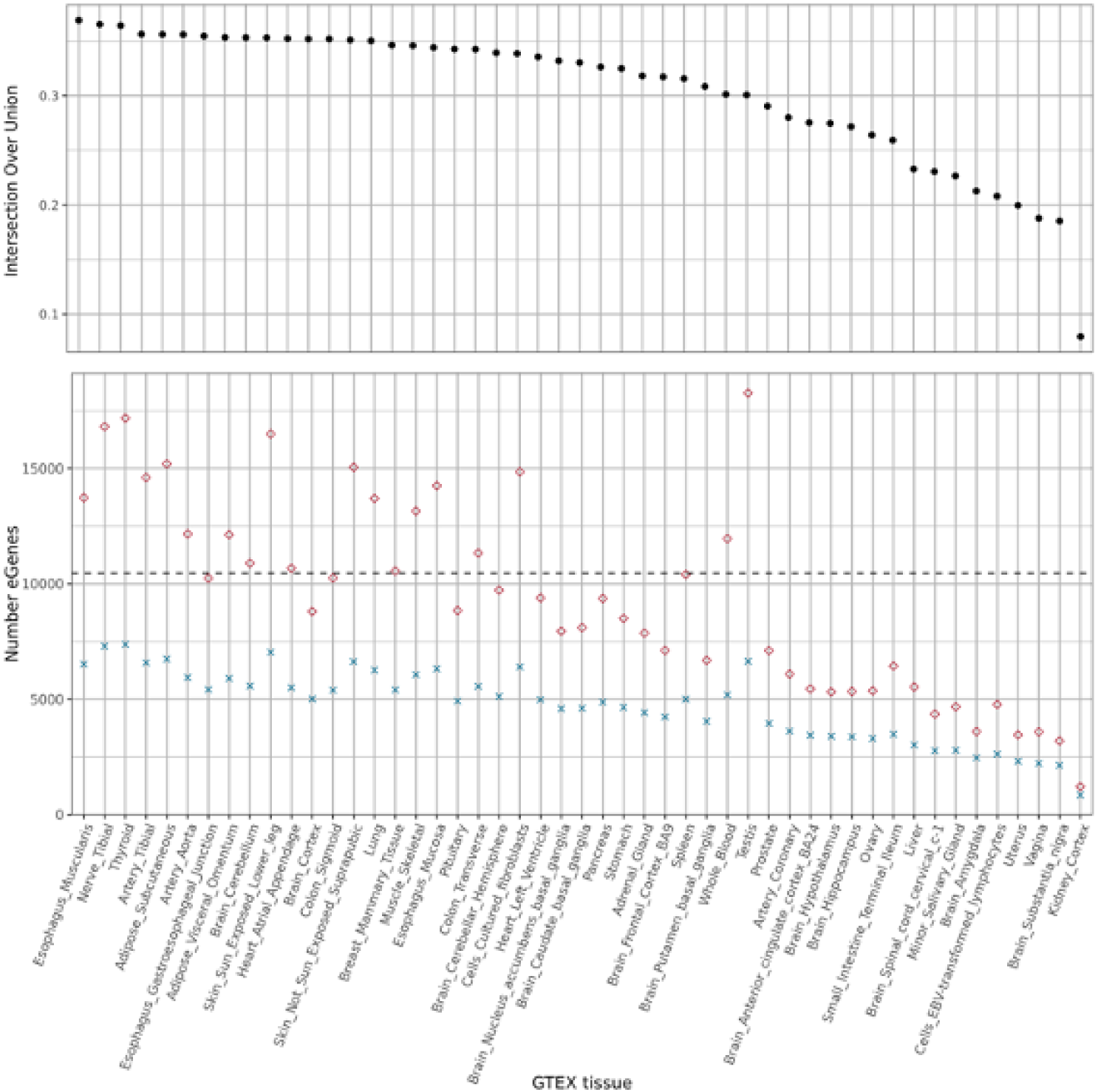
Intersection between eGenes in each GTEX tissue and the METR-eGenes identified in the NSR and/or RPE.

**Supplementary figure 6.**
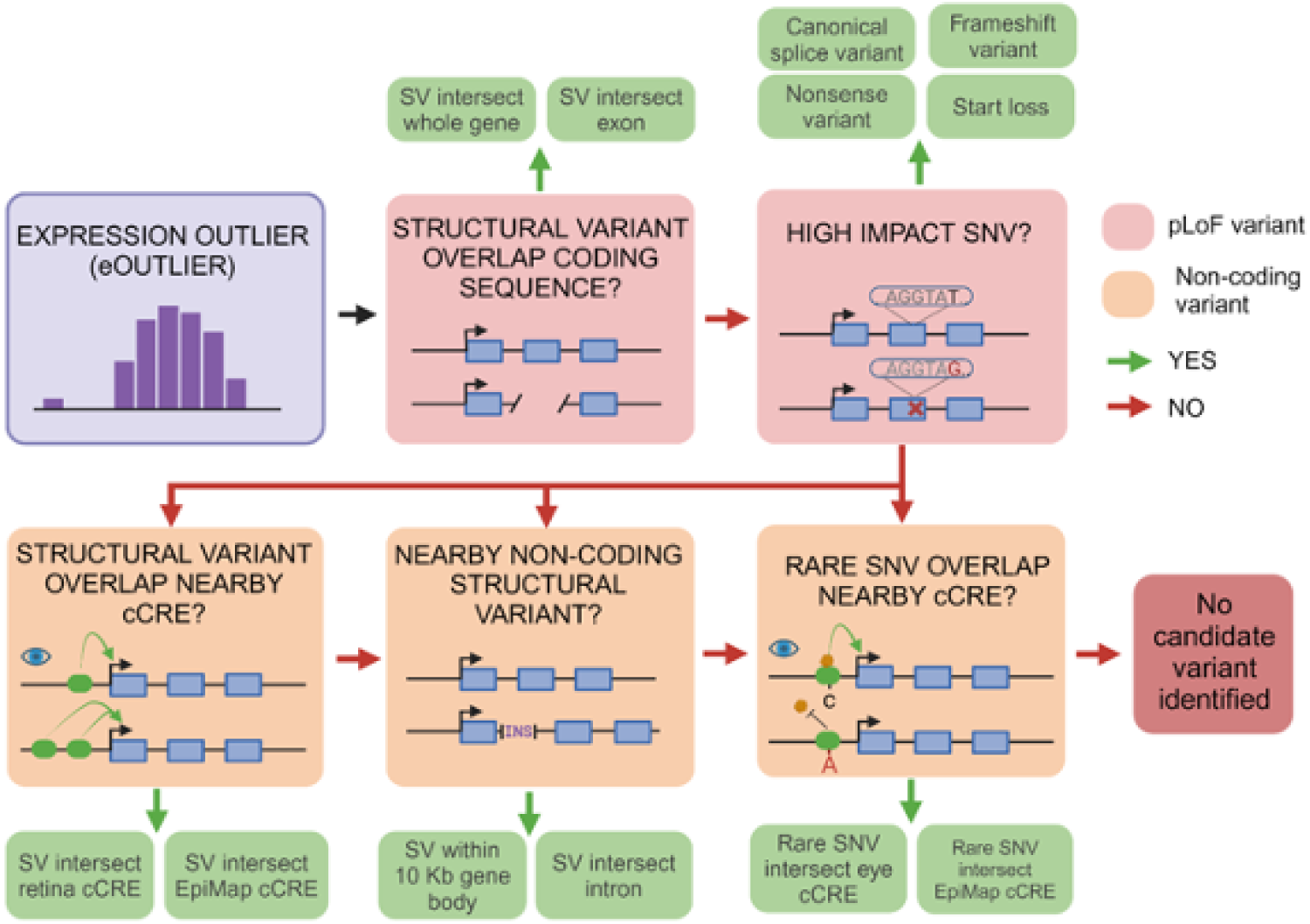
Overview of the hierarchical workflow to identify candidate variants driving outlier expression

## Data Availability

All data produced in the present study are contained in the manuscript or available upon reasonable request to the authors

## Acknowledgments

We express our sincere thanks to the donors, and their families, for enabling this research. This research was supported by the Macular Society (United Kingdom), Fight For Sight, the UK Medical Research Council and the NIHR Manchester Biomedical Research Centre (NIHR203308). This work has also been supported by the Wellcome Trust (224643/Z/21/Z, Clinical Research Career Development Fellowship, P.I.S), the UK National Institute for Health Research (NIHR) Clinical Lecturer Programme (CL-201-06-001, P.I.S), the NIHR Research Professorship grant (RP-2016-07-011, DB) and the NIH/NEI (R01 EY031424 to A.V.S and R01EY035717 to K.M.B.). We thank Selina Mcharg, Nadhim Bayatti, Jay Brown, Simon J. Clark and Paul N. Bishop for the development of the Manchester Eye Tissue Resource. We also thank Stacey Holden, Beverley Anderson and Andy Hayes at the University of Manchester Genomic Technologies Core Facility, and staff at the Ocular Genomics Institute, Harvard Medical School, for their help in the generation of DNA and RNA sequencing datasets for this study. The views expressed are those of the authors and not necessarily those of the funders, including the NIHR and the Department of Health and Social Care.

## Methods

### Gene expression quantification in Neurosensory Retina and Retinal Pigment Epithelium from RNA-Seq data

#### Collection of Donor Eye Tissue

Donor eye tissues were obtained from the Manchester Eye Tissue Repository, an ethically approved Research Tissue Bank (UK NHS Health Research Authority, 15/NW/0932). Eye tissue was acquired after the corneas had been removed for transplantation and explicit consent had been obtained to use the remaining tissue for research. Guidelines established in the Human Tissue Act of 2004 (United Kingdom) and the tenets of the Declaration of Helsinki were adhered to. All ocular tissue was processed within 49 h postmortem. A sample from the iris and ciliary body was collected and stored at −80℃ and the lens and vitreous body were removed. The overlying neurosensory retina was checked for the presence of yellow foveal staining and a macular biopsy was extracted from one eye for further histological analysis. From the other globe, the entire neurosensory retina from the macula and peripheral regions were removed and immediately transferred to RNAlater, to stabilise and protect cellular RNA (Tap et al., 2019). The underlying retinal pigment epithelium (RPE) was mixed with 250 μL phosphate buffered saline (PBS, Merck Millipore) and the cells were gently scraped from Bruch’s membrane, working from the central macular region to the outer peripheral regions. Following centrifugation of the RPE-PBS mixture, the supernatant was removed and RNAlater was added to the RPE pellet and stored at −80℃.

#### RNA sequencing protocol

Total RNA was isolated using an Animal Tissue RNA Purification kit (#25700, Norgen Biotek), as per manufacturer’s instructions. Sequencing was carried out by the Genomic Technologies Core Facility (GTCF) at The University of Manchester. Quality and integrity of the RNA samples were assessed using a 4200 TapeStation (Agilent Technologies) and sequencing libraries were generated using the *Illumina® Stranded mRNA Prep. Ligation* kit (Illumina, Inc.) according to the manufacturer’s protocol. Briefly, total RNA (typically 0.025-1ug) was used as input material from which polyadenylated mRNA was purified using poly-T oligo-attached magnetic beads. Next, mRNA was fragmented under elevated temperature and then reverse transcribed into first strand cDNA. Following removal of the template RNA, second strand cDNA was then synthesized to yield blunt-ended, double-stranded cDNA fragments. Strand specificity was maintained by the incorporation of deoxyuridine triphosphate (dUTP) in place of dTTP to quench the second strand during subsequent amplification. Following a single adenine (A) base addition, adapters with a corresponding, complementary thymine (T) overhang were ligated to the cDNA fragments. Pre-index anchors were then ligated to the ends of the double-stranded cDNA fragments to prepare them for dual indexing. A subsequent PCR amplification step was then used to add the index adapter sequences to create the final cDNA library. The adapter indices enabled the multiplexing of the libraries, which were pooled prior to loading on to the appropriate flow-cell. Paired-end sequencing (2x 75bp) was then performed on an Illumina NovaSeq6000 instrument. Finally, the output data was demultiplexed and BCL-to-Fastq conversion performed using Illumina’s *bcl2fastq* software, version 2.20.0.422. To achieve desired depths of sequencing coverage, NSR samples underwent RNA Sequencing in 5 separate batches while RNA from RPE samples was sequenced in 3 separate batches.

#### Alignment and Expression Quantification

The Genotype-Tissue Expression (GTEx) analysis pipeline (GTEx Consortium, 2019) was followed for quality control of the reads, alignment and expression quantification. All RNA-Seq data was processed uniformly. In summary, alignment of human reference genome GRCh38 was performed using STAR v2.7.4a (Dobin et al., 2013). The GENCODE v38 annotation (Frankish et al., 2019) was used for gene-level expression quantification. Each gene was collapsed to a single transcript using an isoform collapsing procedure designed by GTEX (GTEx Consortium, 2019). RNA-SeQC 1.1.9 (DeLuca et al., 2012) was used for gene-level expression quantification in read counts and transcripts per million (TPM). RSEM v1.3.0 (Li and Dewey, 2011) was used for transcript-level quantifications in transcripts per million. Read counts and TPM values were available for 60,000 genes in both NSR and RPE.

#### Quality control

We assessed RNA-Seq quality control statistics including the total number of reads, number of uniquely mapped reads, number of splice junctions, number of chimeric reads and read length for all NSR and RPE samples. We excluded outlier samples from molQTL mapping (two NSR samples and three RPE samples were removed as read length < 240 bp).

#### Cell type deconvolution from bulk RNA-seq data

We used BayesPrism (Bayesian cell Proportion Reconstruction Inferred using Statistical Marginalization) (Chu et al., 2022) to run a deconvolution model to estimate the proportion of retinal cell types in our bulk RNA-seq data in NSR and RPE. We used single-cell RNAseq data from the ocular posterior segment published by Monavarfeshani et al (2023) as the reference dataset to train the model (study number: SCP2310, downloaded from https://singlecell.broadinstitute.org/single_cell/). The input single-cell dataset for BayesPrism was a cell-by-gene raw count matrix with cell type annotations generated for each cell and the bulk RNAseq dataset was a sample-by-gene raw count matrix. Genes expressed in fewer than 5 cells and those expressed at high magnitude were filtered from the scRNA-seq matrix using the built-in cleanup.genes function. The model was run on protein-coding genes only. The output of the deconvolution model was a set of posterior theta values for each sample in the bulkRNA dataset, corresponding to the estimated proportion of each cell type (from the scRNAseq matrix) that make up the sample.

#### Whole Genome Sequencing Data

Whole genome sequencing of each donor was carried out in 7 separate batches. Five batches were sequenced at the Genomic Technologies Core Facility in Manchester and two batches were sequenced at Ocular Genomic Institute, Massachusetts Eye and Ear Infirmary, Harvard Medical School, Boston. All batches followed the same sequencing protocol and all WGS data was processed uniformly.

Sequencing libraries were generated using on-bead tagmentation chemistry with the Illumina^®^ DNA Prep, (M) Tagmentation Kit (Illumina, Inc.) according to the manufacturer’s protocol. Briefly, bead-linked transposomes were used to mediate the simultaneous fragmentation of gDNA (100-500ng) and the addition of Illumina sequencing primers. Next, reduced-cycle PCR amplification was used to amplify sequencing ready DNA fragments and to add the indices and adapters. Finally, sequencing-ready fragments were washed and pooled prior to paired-end sequencing (2 x 150bp) on an Illumina NovaSeq6000 instrument. Finally, the output data were demultiplexed and BCL-to-Fastq conversion performed using Illumina’s bcl2fastq software, version 2.20.0.422. Genome alignment and variant calling was carried out using Illumina DRAGEN 4.0.3 with Machine Learning and Graph Map Enabled.

#### Aggregated VCF file and variant filtering

Aggregate variant calling was carried out using Illumina DRAGEN 4.0.3 Population Mode. Common variants were used for molQTL mapping (Minor Allele Frequency >2.5% and Minor Allele Count >10).

#### WGS sample-level quality control

We assessed WGS quality control statistics for each sample including median coverage, number of reads with Q>30, percentage genome >15x coverage, uniformity of coverage, total number variants, total number of SNVs, transition/transversion ratio and heterozygous/homozygous ratio. Outlier samples were excluded: one sample was excluded due to high number of variants and high het/hom ratio; and onee another sample was excluded due to high het/hom ratio.

#### WGS variant-level quality control

We applied quality control filters to the aggregate VCF to remove low-quality variant calls using a combination of *bcftools* (v.1.16) and *PLINK* (v.2.0). Firstly, variant calls which did not pass the DRAGEN hard filtering step were removed. Next, we filtered out variant sites which fell within low complexity regions defined by Krusche et al. (2019). Specifically, we removed variant sites which overlapped with all tandem repeats and homopolymers with 5bp slops added on each side (GRCh38_AllTandemRepeatsandHomopolymers_slop5.bed.gz). Subsequently, the remaining variant sites were decomposed and normalised using *bcftools norm* to convert all multiallelic variant sites to biallelic. We then removed variant calls where the inbreeding coefficient was lower than −0.3. Next, genotype calls with GQ<20 and/or where heterozygous calls had allelic imbalance (AB >0.8 or AB <0.2) were set to missing. We also removed monomorphic and compound heterozygous variant calls (ALT1/ALT2), and variants that did not pass the Hardy-Weinberg Equilibrium test (p-value < 10^-8^). Finally, we removed variant calls where missingness was greater than 20%.

#### WGS and RNASeq sample matching

To ensure concordance between paired WGS and RNASeq samples we excluded WGS-RNASeq pairs where the predicted relatedness was <0.8. Relatedness estimates were calculated using Somalier extract (Pedersen et al., 2020). The default polymorphic sites recommended by Somalier were used to infer relatedness. We also assessed the proportion of singleton variants from each WGS sample which were also called from RNASeq data (read depth >15X) for each predicted WGS-RNA pair. Variants were called from RNASeq BAM files using *bcftools mpileup* and the depth at each variant site was calculated using *samtools depth*.

### Molecular phenotypes and covariates for cis-molQTL analysis

#### Formatting and normalisation of RNA-Seq data for expression-QTL analysis

For each tissue, genes which did not meet expression thresholds of >0.1 TPM in at least 20% of samples and ≥6 reads in at least 20% of samples were removed from eQTL analysis. Subsequently, expression values were normalised between samples using the trimmed mean of M-values normalisation (TMM) method (Robinson and Oshlack, 2010) as implemented in edgeR (McCarthy et al., 2012). For each gene, expression values were normalised across samples using an inverse normal transform as implemented by the GTEx analysis pipeline (GTEx Consortium, 2019). A normalised expression matrix was created for each tissue (NSR: 26,734 genes × 183 samples, RPE: 24,448 genes × 176 samples) and used as input for the eQTL analysis.

#### PEER Factors to capture hidden expression confounders

To account for known and unknown biological and experimental confounding factors, a set of 30 covariates were generated for each RNA-Seq sample using the Probabilistic Estimation of Expression Residuals (PEER) method (Stegle et al., 2010) applied to normalised gene expression levels. The PEER method is based on a Bayesian network which assumes that expression levels are influenced by additive effects from multiple independent sources (Stegle et al., 2010).

#### Genotyping Principal Components

Principal component analysis with EIGENSOFT 6.0.1 (Patterson, Price and Reich, 2006) was carried out to capture ancestral variation within the cohort. The top five principal components for each participant were used as covariates in the eQTL analysis.

#### eQTL mapping with tensorqtl

TensorQTL (Taylor-Weiner et al., 2019) was used to find genetic variants which were significantly associated with the expression of nearby genes (up to 1 Mb away) in NSR and RPE. The required input files were the normalised gene expression matrix, the binary and filtered genotype data and a covariates table which included the following information for each participant: sex, WGS batch, five top principal components and 30 PEER factors. TensorQTL generates nominal p-values for each variant-gene pair based on a linear regression model between genotype and expression. Additionally, by using a permutation model, it can identify the variant which is most significantly associated with the expression of each gene.

#### Linear regression model for nominal gene-variant associations

For every input gene in NSR (n = 26,734 genes) and RPE (n = 24,448 genes), the following linear regression model was tested by TensorQTL for each variant within a 1 Mb-window (upstream and downstream) of its transcription start site:

Gene expression ∼ Variant genotype + Sex + Five top Principal Components + 30 PEER Factors + WGS Sequencing Batch

Gene expression was assigned as the dependent variable and the variant genotype as the predictor of interest. However, gene expression quantification is dependent on multiple biological and technical factors. Therefore, additional covariates are included in the regression model. Nominal p-values were generated for each variant-gene pair by testing the alternative hypothesis that the effect size (or slope) of the variant genotype deviates from 0. To account for false positives, a false discovery rate (FDR) threshold was also calculated for each gene using a permutation scheme.

#### Permutation scheme to find candidate eQTL for each gene and False Discovery Rate threshold

The aim of the permutation scheme was to find the best nominal association for each gene and assess its global significance (Ongen et al., 2016). TensorQTL implements a beta-approximation model to calculate the number of permutations for each gene (Taylor-Weiner et al., 2019). This model is based on the hypothesis that P-values obtained through permutations are beta-distributed (Dudbridge and Koeleman, 2004). The beta-approximation model is based on estimating the shape parameters for the beta distribution of gene-variant p-values for each gene by maximum likelihood (Galwey, 2009). This estimation is calculated after generating a null set of P-values for each gene from a random number of permutations. The number of permutations required to generate the null set of P-values is lower than the number of permutations required to empirically calculate the beta distribution shape parameters, thus reducing computational burden. Subsequently, the smallest nominal p-value can be assigned an adjusted beta-p-value.

Moreover, the adjusted beta p-values are used by TensorQTL to calculate q-values using the Storey and Tibshirani False-Discovery Rate (FDR) procedure (2003). Genes with a significant eQTL (eGenes) are those where FDR ≤0.05 for the most significant QTL association. To identify all significant variant-gene pairs for each eGene, a nominal p-value threshold (***p_t_***) was set based on the beta distribution model obtained from the permutations for that particular eGene (Ongen et al., 2016). Therefore, a significant association between an eGene and a nearby genetic variant (eVariant) is one where the nominal p-value ≤p_t_, which corresponds to FDR ≤0.05. The genetic loci that are associated with the expression of a nearby eGene are referred to as cis-expression Quantitative Trait Loci (cis-eQTLs).

#### Allelic fold change

To quantify the eQTL effect size we estimated the log allelic fold change (aFC), following the method established by Mohammadi et al. (2017). The aFC is defined as the log-ratio between gene expression on the same chromosome as the eVariant ALT allele and the expression on the same chromosome as the REF allele.

### Comparison with other eQTL studies

#### Intersection with GTEX eQTLs

All significant eQTL associations were downloaded from the GTEx Open Access portal (v8) for each available tissue (https://www.gtexportal.org/home/downloads/adult-gtex/qtl). We calculated the intersection between the number of METR-eQTLs which were also shared by each GTEx tissue. For an eQTL to be considered shared between both datasets, the variant ID (*CHR_POS_REF_ALT*, with GRCh38 coordinates) and the Ensembl gene ID (without version number) had to match. We compared the similarity between each GTEx tissue and the METR-eQTL dataset using the Intersection over Union (IoU) statistic. The IoU is the ratio of the number of eQTLs present in both sets over the total number of eQTLs in one set and/or the other. Therefore, the IoU statistic considers the different number of eQTLs present in each GTEx tissue. We also calculated the number of shared eGenes between each GTEx tissue and the METR-eQTL dataset, as well as the degree of eGene similarity using the IoU statistic.

#### Intersection with EyeGEx and the Eye eQTL Atlas

We compared all METR eQTLs with retina eQTLs mapped by EyeGEx (Ratnapriya et al., 2019). METR variant IDs were converted to rsIDs using dbSNP build release 156 (Phan et al., 2025). First, we identified genes had been associated with eQTLs in EyeGEx and in our study (common eGenes). For these genes, we extracted the top eQTLs identified by EyeGEx and checked if they were replicated in our cohort or if they were in high LD (r2 > 0.8) with a METR-NSR eQTL. Pairwise LD scores were calculated using LDlinkR (Myers et al., 2020).

Additionally, we queried the METR-eQTL dataset to understand if we replicated eQTLs that had been identified through TWAS and colocalization analyses as candidate variants which impact AMD risk genes (Orozco et al., 2020; Ratnapriya et al., 2019). We also tested if the candidate variants were in high LD with METR-eQTLs or if novel METR-eQTLs had been identified for the risk gene. We downloaded eQTLs from Orozco et al. (2020) from the eye eqtl browser, available at (https://eye-eqtl.com/).

#### Annotation of eVariants and bootstrapping analysis to calculate enrichment of eQTLs in characterised regulatory loci

All NSR and RPE eQTL variants were annotated with the Ensembl Variant Effect Predictor (McLaren et al., 2016). We also annotated all eVariants with cell-type agnostic candidate cis-regulatory element (cCRE) annotations from the ENCODE Encyclopaedia of cCREs (Snyder et al., 2020), which included promoters, proximal/distal enhancers, CTCF-only binding sites and DNase-H3K4me3 sites (poised elements). To include tissue-specific cCRE annotations, we annotated eVariants with characterised regulatory loci from retina, RPE and macula from Cherry et al. (2020). Additionally, we annotated variants with non-eye specific cCREs from adult tissues in EpiMap (Boix et al., 2021). Specifically, we downloaded all enhancer and promoter BED files from all adult tissue types from https://compbio.mit.edu/epimap/. We excluded cCREs from embryonic and neonatal cell types, and all primary cell lines as their regulatory patterns may be different from adult human tissues.

To calculate the relative enrichment of eVariants which overlapped with each type of regulatory element, we used bootstrapping analysis. We carried out 1,000 subsampling iterations. For each iteration, 100,000 eVariants were randomly selected with replacement. We then matched each eVariant with another non-eQTL variant from our cohort that had a similar allele frequency and gene density (number of gene TSSs within 1Mb of the variant). Each subset of eVariants and matched control variants was intersected with the cCRE datasets described above using BEDtools (Quinlan and Hall, 2010). We then calculated the relative enrichment as the ratio of eVariants to control variants which overlapped each type of cCRE.

For each cCRE type we then calculated the mean relative enrichment and the 95% confidence interval. If there was no relative enrichment, we would expect the mean score to equal 1. Therefore, to calculate significance p-values from the bootstrapped results we created a null distribution where *mean = 1* and *std. deviation = std. deviation of the bootstrapped result set*. We then calculated the Z-score between the observed bootstrapped mean and the mean of the null distribution.

#### Identification of transcriptome outliers using the DROP pipeline

We utilised the DROP v.1.4.0 snakemake pipeline to identify transcriptome outliers from NSR and RPE. We ran the aberrant expression module with the parameters listed in *Methods Table 1* to identify expression outliers. The aberrant splicing and monoallelic expression modules were applied to NSR samples with the parameters listed in *Methods Table 2* and *Methods Table 3* respectively.

**Methods Table 3.** Parameters used to run the aberrant expression module on DROP 1.4.0.

| Parameter | Value |
| --- | --- |
| Groups | NSR, RPE |
| FPKM Cutoff | 1 |
| Implementation | Autoencoder |
| Z-score cutoff | 0 |
| p-adj cutoff | 0.05 |
| Max tested dimension proportion | 3 |
| Yield size | 2,000,000 |

**Methods Table 4.** Parameters used to run the aberrant splicing module on DROP 1.4.0.

| Parameter | Value |
| --- | --- |
| Groups | NSR |
| recount | FALSE |
| longRead | FALSE |
| keepNonStandardChrs | FALSE |
| filter | TRUE |
| minExpressionInOneSample | 20 |
| quantileMinExpression | 10 |
| minDeltaPsi | 0.05 |
| FRASER_version | FRASER2 |
| deltaPsiCutoff | 0.1 |
| quantileForFiltering | 0.75 |
| Implementation | PCA |
| p-adj cutoff | 0.05 |
| Max tested dimension proportion | 6 |

**Methods Table 5.** Parameters used to run the monoallelic expression module on DROP v.1.4.0.

| Parameter | Value |
| --- | --- |
| Groups | NSR |
| gatkIgnoreHeaderCheck | TRUE |
| padjCutoff | 0.05 |
| allelicRatioCutoff | 0.8 |
| addAF | TRUE |
| maxAF | 0.001 |
| maxVarFreqCohort | 0.05 |

#### Hierarchical workflow to identify candidate variants driving outlier expression

We designed a hierarchical workflow to identify candidate variants driving outlier expression (eOutliers) using snakemake version 7.32 (*Methods Figure 1)*. Each eOutlier was identified by a unique sampleID-geneID combination. Briefly, the workflow would first identify a pLoF variant from the eOutlier sample in the corresponding eOutlier gene, which could be an exonic structural variant, or a SNV with a high impact consequence based on *Ensembl’s Variant Effect Predictor (v*.*112.0).* High impact SNVs included nonsense, frameshift, start/stop loss and canonical splice donor/acceptor variants. If no pLoF variant could be identified, the workflow would then search for regulatory variants which were within 10Kb of the eOutlier gene body. Regulatory variants were defined as structural variants and rare SNVs which overlapped with nearby retina cCREs or non-retina specific cCREs from different adult tissues in EpiMap. If no regulatory variant was identified, the model would check if any other non-coding structural variant fell within 10Kb of the eOutlier gene body, before returning a negative search result. All intersection steps were carried out using *bedtools intersect.* We used *bcftools* to extract SNVs from each outlier sample for the corresponding eOutlier gene from the filtered aggregate VCF. Rare SNVs were defined as those where the gnomad v4 allele frequency was lower than 1%.

The output of the workflow is a JSON object, where each eOutlier ID is assigned a category based on the candidate variant type (*Methods Table 4)*, alongside details of the eOutlier metrics from OUTRIDER and a list of the candidate variant/s for further analysis.

**Methods Table 6.** Output categories for each eOutlier based on the identified candidate variants driving outlier expression.

| eOutlier category based on the candidate variant | Variant type |
| --- | --- |
| SV_CNV_intersect_whole_gene | pLoF variant |
| SV_CNV_intersect_exon | pLoF variant |
| High_impact_SNV_disrupt_gene | pLoF variant |
| SV_CNV_intersect_Eye_cCRE | Regulatory variant |
| Rare_SNV_Intersect_Eye_cCRE | Regulatory variant |
| SV_CNV_intersect_EpiMap_cCRE | Regulatory variant |
| Rare_SNV_Intersect_EpiMap_cCRE | Regulatory variant |
| SV_CNV_intersect_intron | Non-coding structural variant |
| SV_CNV_within_10Kb_gene_body | Non-coding structural variant |
| Common_SNV_Intersect_cCRE | No candidate variant |
| No_SV_or_SNV_of_interest | No candidate variant |

**Methods Figure 1.**
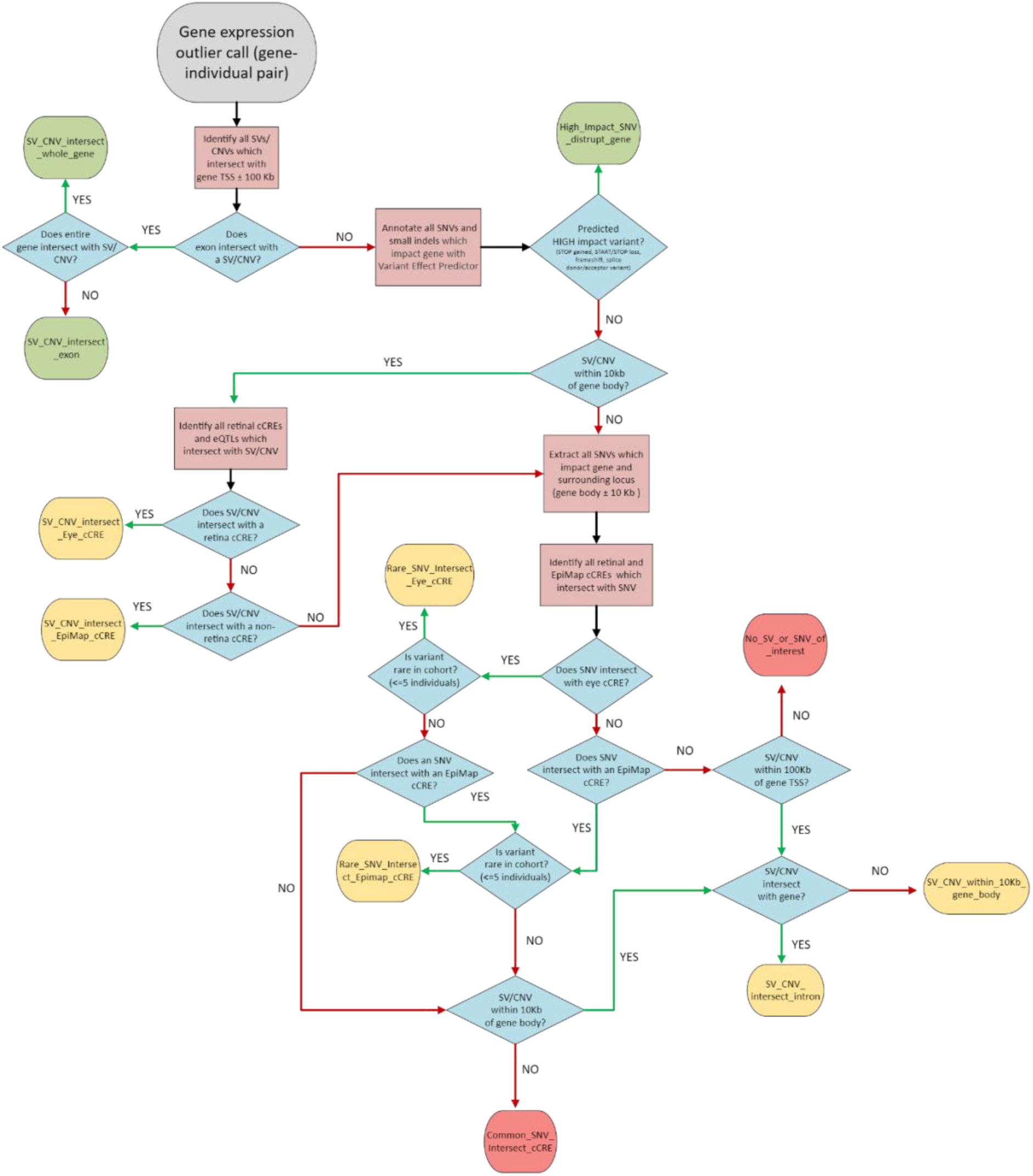
Workflow used to identify candidate variants driving eOutlier events. The workflow was automated using snakemake, where each processing step was defined by an individual rule, and decision branches were defined using checkpoints.

#### Implementation of Watershed

For all genes with an eOutlier in the NSR (n = 702), we extracted all rare variants (gnomAD allele frequency < 1%) which intersected with the gene body ± 10Kb. Variants were extracted for all samples with NSR RNASeq data (n = 183) from the post-QC aggregate VCF. We then annotated all rare variants with selected annotations from VEP (McLaren et al., 2016) and CADD (Schubach et al., 2024) *(Supplementary Table 6)* and intersected them with known retina-specific cCREs from Cherry et al. (2020) and non-retina specific cCREs from EpiMap. Missing annotations were replaced with default imputation values obtained from the CADD release notes (*Supplementary Table 6)*.

The scripts required to run the Watershed model were downloaded from https://github.com/BennyStrobes/Watershed. Each line of the input dataset corresponded to a single gene-sample pair and included a set of annotations describing the rare variants nearby the gene and the p-values corresponding to the aberrant expression, aberrant splicing and monoallelic expression modules from DROP for NSR and RPE. From this data, the model was able to optimise the appropriate parameters using Watershed Exact optimisation and then calculate the posterior probabilities that a particular set of genomic annotations would lead to each significant eOutlier p-value (adjusted p-value < 0.05).

For each gene-sample, Watershed required a single value for each annotation listed on *Supplementary Table 6*, even though more than one rare variant may lie within the gene of interest. Therefore, from the set of annotated rare variants corresponding to a particular gene-sample, we selected the annotation which was considered the most informative (generally the maximum value). Therefore, each line of the input dataset for Watershed described the landscape of rare variants which surrounded a particular gene. Watershed also required N2 pairs, or pairs of individuals which shared the same rare variant to evaluate the model. We randomly selected 3000 N2 pairs from our dataset.

The Watershed model was run using the predict_watershed.R script with an adjusted p-val threshold of 0.05 and the number of dimensions set to 6 (corresponding to the 3 DROP p-values from NSR and 3 from RPE).

